# Strategies and interventions to strengthen pharmacovigilance systems in low- and middle-income countries: a scoping review

**DOI:** 10.1101/2022.12.06.22283123

**Authors:** Olga Menang, Andrea de Soyres-Kuemmerle, Karen Maigetter, Christian Burri

**Author notes:** Corresponding to Olga Menang.

## Abstract

**Introduction:** In the past decades, numerous strategies have been proposed to strengthen pharmacovigilance (PV) systems in low-and-middle-income countries (LMIC). Though there has been progress, many PV systems in LMIC are still not performing adequately. The objectives of this scoping review are to describe strategies to strengthen PV in LMIC and to propose recommendations for future investments in capacity building.

**Methods:** The review was conducted following the Joanna Briggs Institute (JBI) guidelines on conducting scoping reviews. Literature searches were performed in MEDLINE, EMBASE, Web of Science, PDQ-evidence, CINAHL and relevant websites from 1990 to January 2021. Publications included were primary studies, articles, and policy and guideline papers, describing interventions to strengthen PV in LMIC. Two reviewers independently screened titles, abstracts and full-texts, and one reviewer performed data extraction and carried out a descriptive data analysis.

**Results:** 10,903 unique titles were screened and 152 were eligible for full text review. Of these, 57 and an additional 13 reports from grey literature fulfilled eligibility criteria for inclusion in the review. Forty-five papers (64%) described interventions aimed at increasing PV knowledge and reporting of adverse drug reactions (ADR), primarily education of healthcare professionals (HCP), alone or in combination with other interventions such as mobile and electronic reporting, enhanced passive and active surveillance. Twenty-five papers (36%) discussed interventions or strategies implemented at the national targeting different components of the national PV system, such as legal basis and guidelines for PV, PV organisation and infrastructure and PV procedures.

**Conclusions:** Results of this review suggest that educating HCP on ADR reporting is the most common approach to build PV capacity in LMIC. Though important, education alone is insufficient and should ideally be organised within the holistic framework of strengthening national PV systems, with a focus on building capacity for advanced activities such as signal detection.

**Key message:** There are numerous strategies and recommendations to strengthen PV systems in LMIC. However, the effective implementation of these strategies has not been extensively described in literature. This scoping review describes different interventions and strategies that have been implemented to build and improve PV capacity in systems in LMIC. An overview of current PV strengthening strategies and interventions, and when available, their outcome and impact, is useful to guide decision making for future investments in PV development in developing countries.

**Trial registration:** Open Science Framework (https://osf.io/ge56p)

## INTRODUCTION

Several low-and-middle-income countries (LMIC) are projected to receive novel vaccines and medicines in the coming years, some of which target diseases endemic in LMIC, e.g., malaria, dengue and Ebola. Therefore, pharmacovigilance (PV) systems in these countries must be efficient and functional, in order to effectively monitor the safety and effectiveness of these novel products. In the past three decades, several global and local initiatives have aimed at building PV capacity in LMIC. Earlier assessments of PV systems in LMIC showed that only few countries had performing PV systems that could detect, evaluate, and prevent medicine safety issues (1, 2). Though more recent evaluations show major improvement, (3, 4), a majority of national regulatory authorities (NRA) in LMIC have not yet attained World Health Organisation (WHO) Global Benchmarking Tool (GBT) performance maturity level 3 (stable, well-functioning and integrated regulatory system) and level 4 (regulatory systems operating at advanced level of performance and continuous improvement) (5).

Numerous initiatives have aimed to strengthen PV in LMIC in the past decades. In 2011, the WHO Global Vaccine Safety Blueprint (GVSB 1.0) proposed a three-part strategy for building vaccine PV: i) build minimum capacity for passive vaccine safety surveillance in all countries, ii) build enhanced capacity for active surveillance in countries where newly developed vaccines will be introduced, and iii) establish a global vaccine safety support structure (6). In 2013, a five-part PV enhancement strategy was proposed in a report submitted to the Safety and Surveillance Working Group of the Bill and Melinda Gates Foundation (BMGF): i) focus on a global health product pipeline, ii) implement a risk-based prioritization of candidate drugs and vaccines, iii) invert the current capacity building paradigm, iv) incorporate sustainability from outset, and v) plan scalability (2). In 2021, the WHO Global Vaccine Safety Blueprint 2.0 (GVSB2.0) proposed to move from minimal and enhanced capacity concept proposed in GVSB1.0 (6) to maturity levels, using a scale of 1 (least developed) to 4 (regulatory system operating at advanced level), to assess the level of development of monitoring and regulatory systems (7).

The slow progress of PV in LMIC not only raises questions about core challenges on the growth of PV, but also about the appropriateness of strategies and interventions used so far to develop PV. Describing these strategies and if possible, their outcomes would be useful to guide decision making for future national PV strengthening initiatives. There have been only few scoping and systematic reviews on topics related to PV globally. These include reviews on interventions to improve adverse drug reactions (ADRs) reporting among health care professionals (HCP) (8, 9) and the use of social media for PV (10, 11). However, these reviews were not solely focused on LMIC and did not describe a holistic approach to developing PV in LMIC. The objectives of this current scoping review are to describe strategies and interventions that have been implemented to strengthen national PV systems in LMIC and to recommend areas for future investments in capacity building. As PV systems in LMIC continue to mature, supported by multilateral agencies and donors, synthesised information on the different enhancement approaches will be useful for national and global PV stakeholders, including donor agencies, as they continue to build effective and sustainable PV systems.

## METHODS

### Research questions

1. What interventions or strategies have been used to build and strengthen national PV systems in LMIC in the last three decades?
2. What is the impact of these strategies and what recommendations can be made to policy-makers for implementing and/or strengthening PV systems in LMIC?

### Study design

The scoping review was conducted in accordance with the Joanna Briggs Institute (JBI) methodology for scoping reviews (12) and conformed with the PRISMA-ScR (Preferred Reporting Items for Systematic Reviews and Meta-Analyses Extension for Scoping Reviews) (13). The protocol was developed according to the JBI scoping review template and was registered with Open Science Framework.

### Eligibility criteria

Publications or papers within the last 30 years (1990 to 2021) that described strategies or intervention to establish or strengthen national PV systems in LMIC were included. Also included, were reports describing PV systems if these included interventions to develop PV. Lastly, national PV guidelines, policy and strategic documents from NRA were also included.

### Information sources

This scoping review included primary research, both qualitative and quantitative, that described interventions or strategies to strengthen PV capacity, as well as policy and guideline papers from NRA. Literature searches were performed in MEDLINE, EMBASE, Web of Science, CINAHL and PDQ-Evidence. Sources of grey literature included Google Scholar, websites of technical agencies such as WHO and websites of selected NRA. A selection of NRA websites was necessary, as it was practically impossible to search all LMIC NRA websites. Therefore, websites of countries selected to participate in the next phase of the research (semi-structured interview on the same research question) were searched.

### Search strategy

An initial limited search of MEDLINE, EMBASE, Epistemonikos, JBI Database of Systematic Reviews and Implementation Reports and Web of Science was undertaken to identify articles of interest. A librarian used text words contained in the titles and abstracts of relevant articles and the index terms used to describe these articles, to develop a full search strategy in MEDLINE (Appendix I). This search strategy was adapted for each database cited above using the Polyglot Search Translator (14). A simpler search strategy was used to search Google Scholar. A grey literature search was conducted according to the Canadian Agency for Drugs and Technologies in Health Guide (15) (Appendix 2).

### Study selection

Identified citations from the searches were collated and uploaded into EndNote X9.3 (Clarivate Analytics, PA, USA) and duplicates removed. To facilitate study selection, a decision tree was developed and tested by two reviewers, [Olga Menang (OM) and Andrea de Soyres-Kuemmerle (ASK)], on a random sample of 100 titles and abstracts. Three batches of 100 titles and abstracts were tested until both reviewers agreed on the selection. OM reviewed 100% of titles and abstracts while ASK reviewed 15%, as selected titles and abstracts were similar for both reviewers for the first 1500 titles and abstracts screened. Two batches of 10 full-text articles were tested until agreement on article selection; both reviewers reviewed all full texts selected at title and abstract screening. The results of the search and the study inclusion process were reported in full and presented in a Preferred Reporting Items for Systematic Reviews and Meta-analyses extension for scoping review (PRISMA-ScR) flow diagram (13).

### Data extraction

Data were extracted using a data extraction tool developed for the purpose by the reviewers (Appendix III). Extraction parameters included the nature and description of the intervention, the outcome, the challenges encountered and the lessons learnt. Extracted data was reviewed by ASK and a third reviewer, Karen Maigetter (KM), and was revised as needed.

### Data analysis and presentation

The extracted data were presented in a descriptive summary aligned with the objectives of the review. Data was analysed by strategy, nature and types of intervention to strengthen PV systems, and if the strategy or interventions targeted one or several components of the PV system. Risk of bias or quality appraisal across studies was not conducted, which is consistent with the Joanna Briggs Institute Methods Manual, because the scoping review method is not intended to be used to critically appraise (or appraise the risk of bias of) a cumulative body of evidence (13).

## RESULTS

### Study selection

The study selection flow diagram is presented in Figure 1. 10,903 unique titles were screened, of which 152 articles were selected during title and abstract screening. Of these 152 articles, 57 articles and an additional 13 articles from Google Scholar and grey literature searches fulfilled eligibility criteria for inclusion in the review (Figure 1). Appendix VI provides citation details of papers excluded on full-text examination and reasons for their exclusion.

**Figure 1:**
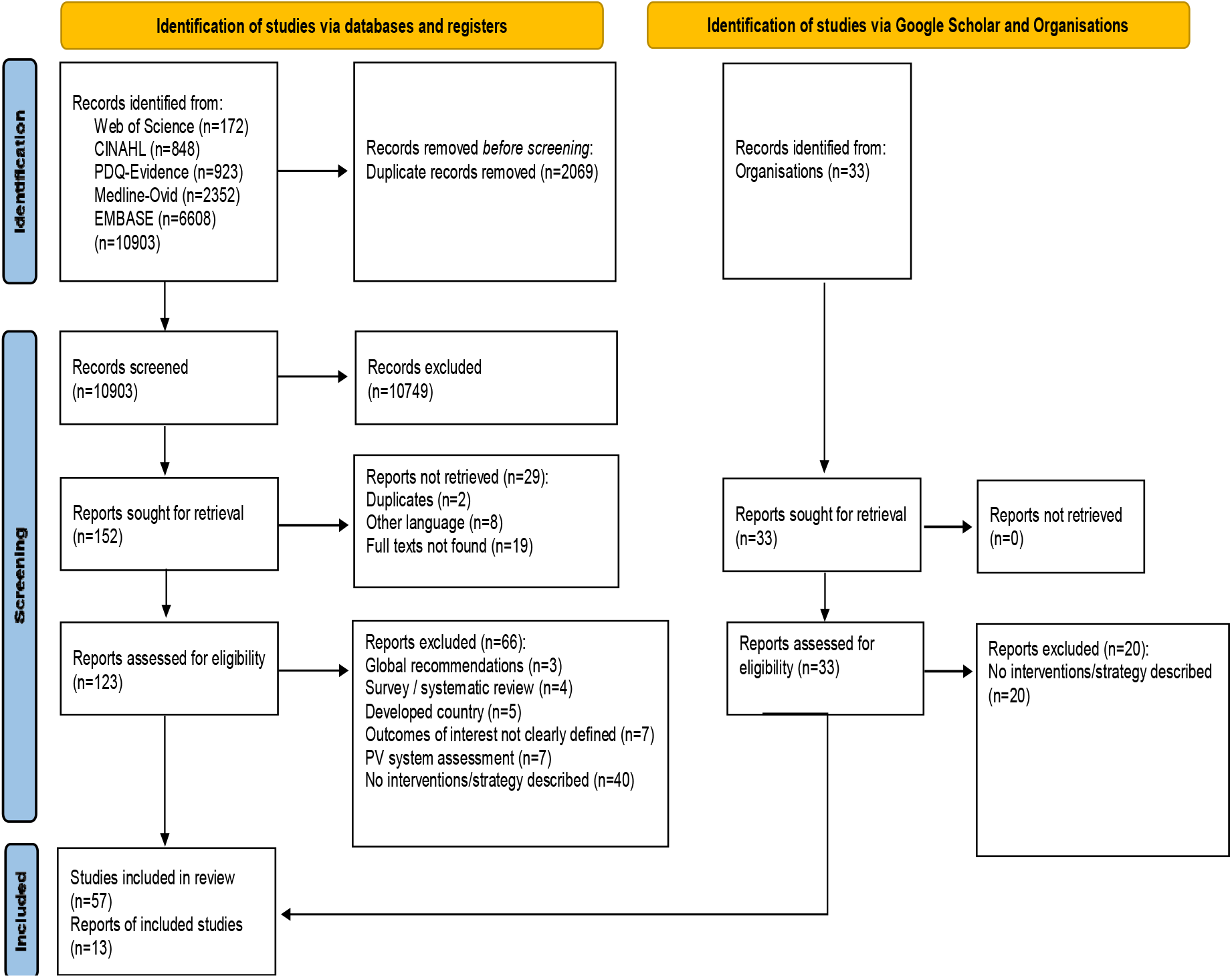
Study selection flow diagram.

### Nature of interventions and strategies

The regions and countries of papers included in the analysis of this review are presented in Appendix IV. Papers from East Asia Pacific region (9), Europe and central Asia (1), Latin America and the Caribbean (6), Middle East and North Africa (5), South Asia (17), Sub-Saharan Africa (28) and 5 papers providing global/LMIC recommendations were included. Strategies and interventions identified to strengthen PV systems in LMIC were divided into 2 categories: 1) Interventions aimed at increasing PV knowledge and reporting of adverse events (AEs) [ADRs or adverse events following immunisation (AEFI)] and, 2) Interventions aimed at building or strengthening various components of the national PV system. Details of each intervention are provided in Appendix V.

### Interventions aimed at increasing PV knowledge and AE reporting

Forty-five papers described interventions aimed at increasing HCP PV knowledge and reporting of ADRs or AEFI. These included educational interventions (lectures or workshops), alone or in combination with other interventions such as mobile and electronic reporting, enhanced passive and active surveillance or providing financial incentives.

#### Educational interventions

Although all publications included in the analysis had an educational component, 21 out of 70 (30%) reported exclusively educational or training workshops, aimed at educating and sensitizing HCP on AE reporting and PV. Primarily provided by health institutions, these interventions were not reported as part of a broader PV strengthening scheme. The format, content, and duration varied, with duration ranging from 45 minutes (16) to several days (17). For a majority of these interventions, the content was information on ADR reporting and raising awareness on PV. In some cases, educational material such as handouts and job aids were distributed. Some interventions included practical sessions on completing AE reporting forms (18), feedback or reminders to HCP (19), and follow-up contacts to assess changes in AE reporting (20) or to address any challenges encountered (21).

There was an increase in the knowledge, attitude and practice skills of PV and ADR reporting rates for most educational interventions. After interventions, some papers reported a 12-fold increase in the number of ADRs reported and an increase in the quality of ADRs (22), or a fourteen-fold increase in ADR reporting (19), or as high as a 70-fold increase in the number of ADR reports (18). Only very few papers reported a small impact of education training. One of these was the training of naïve HIV-positive patients with limited literacy to identify and report ADRs using pictorial representation (23).

#### Mixed interventions

Eleven papers reported education in combination with other interventions such as financial incentives and changes in institutional ADR reporting policies. Two papers described provision of financial compensation (in addition to other changes) as an incentive for reporting ADRs in a hospital setting. For the first, ADRs increased from 29 at pre-intervention, to 277 at first intervention, to 666 at second intervention period, with an increase in the quality of reports (24). For the second, ADRs increased from 577 and to 832 after intervention, with an increase in reporting of serious ADRs and improvements in reporting compliance (25).

Other inventions included follow-up visits (26), providing feedback to HCP on ADR reporting and reviewing the quality and completeness of ADRs (27) or distributing standard operating procedures on ADR reporting, improving reporting mechanisms and supervisory visits (28). In Mozambique, in addition to training workshops on PV, a ‘yellow card’ system for spontaneous reporting of ADRs was implemented, a district focal person to facilitate communication between the health staff and the PV unit was designated and after 5 months, HCP were retrained to reinforce learnings from the first workshop (29). These papers also reported an increase in PV knowledge and number of ADRs reported.

#### Mobile phone and electronic reporting

Seven papers in this review described the use of mobile phones, mobile applications and electronic reporting platforms as mechanisms to increase AE reporting. For example, in Rwanda, patients were educated to report ADRs using mobile phones either by accepting a call from physicians or by calling physicians directly. Compared to the previous months where no ADRs were reported, 34 patients reported an ADR after intervention (30). Likewise, in Cameroon, 20 AEFI were received from a group reporting with a mobile phone, compared to one AEFI from the group reporting physically at the health facility (31). Other papers reported the development of mobile applications for reporting ADRs reporting (32) and supporting ADR evaluation and causality assessment(33).

At the national level, in Brazil, the national electronic reporting system, Notivisa, was replaced by VigiFlow™ and implemented at all levels of the health systems including a module for the public and another for industry. HCP were trained on PV during the process and the new tool was promoted through different platforms, resulting in a 62% increase in ADRs received by Anvisa compared to the same period for the previous year, including reports from the public (34). Likewise, Mexico implemented VigiFlow™, and e-reporting throughout the country, enabling ADR reporting from health services and industry, as well as analysis of safety data (35).

#### Passive and active surveillance

Six papers in the current review reported implementation of enhanced passive and/or active surveillance as mechanisms to increase reporting and collect more reliable PV data. These papers reported an increase in the number, timeliness and quality of ADRs collected (36-38) even though one paper reported that the numbers reduced in subsequent months (39).

### Interventions aimed at strengthening various components of the national PV system

Twenty-five papers discussed interventions or strategies implemented at the national level that aimed to strengthen different components of the PV system. For example, PV in India was implemented in a stepwise approach, targeting different components and stakeholders of the PV systems. These steps included: i) development and revision of PV guidelines; ii) establishing and training national safety and AEFI committees; iii) setting up regional ADR monitoring centres; vi) developing communication tools and guidelines; v) integrating PV in public health programs (PHP); vi) android mobile application and toll-free helpline to enable reporting of ADRs; and vii) safety communication to stakeholders and the public though different media (40-42). A similar comprehensive approach was reported in Vietnam, and within 7 years of the creation of the national PV centre, both the quality and the quantity of ADRs improved (43).

In Malawi, the national PV system was enhanced through a combination of strategies targeting the PV organisation and infrastructure, PV procedures and AE reporting, national coordination of PV activities and training of PV personnel and HCP. During the 18 months of implementation, 443 HCP at 61 healthcare facilities were trained in PV and the ADRs and AEFI increased from 22 at pre-intervention to 228 (44). Other interventions identified at the national level included creation of the national PV centre, including PV in curriculum of medicine, pharmacy, and dentistry schools, and building collaborations between NRA and PHP (45), or development of PV policies and tools; establishment and training national safety review committees; and establishment of country coordination teams (46).

In 2019, the National Drug Authority of Uganda published a 5-year (2018 to 2022) strategic plan to develop PV. The plan defined four strategic areas of development: i) build PV infrastructure and technical capacity; ii) PV policy enhancement; iii) collaboration and information exchange; and iv) visibility and awareness of PV and safety related information (47). In 2020, the Nigerian National Pharmacovigilance Policy was revised, providing a framework for the implementation of the policy, covering the entire scope of PV, including stakeholders, institutionalisation of PV policy documents, PV structures, human resources, industry, funding and monitoring and evaluation (48). In Namibia, results of a qualitative survey recommended several strategies on the integration of PV systems in public health: i) enhancing reporting by integrating PV into the daily practice of HCP; ii) establishing policies addressing PV activities; iii) capacity-building through continuing professional education and support, advocacy, stakeholder engagement; iv) incentivising and recognition for PV performance; and v) facility-based policies for universal and inclusive reporting (49).

Also identified in this review were global recommendations on strengthening national PV systems. These included WHO’s Project 3-S (Smart Safety Surveillance), aimed at optimizing post marketing surveillance of priority medicines and vaccines in LMIC. Six core strategies were proposed: i) adopt a stepwise approach with an initial pilot for three new products; ii) leverage available resources from WHO partners and national PV centres; iii) partnership with industry; iv) develop a holistic country plan for PV as part of medicines regulation; v) collaboration with other global initiatives; and vi) build PV infrastructure progressively, from minimum to mid-range and advanced capacity (50). In 2018, Elshafie et al., proposed different strategies to encourage the introduction and sustain the advancement of robust PV systems in developing countries: i) enhancing spontaneous reporting though a combination of interventions; ii) training HCP and incorporation PV into educational institutions and curricula of HCP; iii) actively engaging the public to participate in PV; iv) facilitating ADR reporting by simplifying the process and enabling mobile and electronic reporting and enabling adequate data management and communication of safety signals; v) establish well-organized healthcare systems, with effective policies and strict drug regulations; and vi) implement regulations and guidelines for industry (51). Lastly, in 2021, PhArmacoVigilance Africa (PAVIA) released the “Guide for Effective Implementation of the Pharmacovigilance Policy in Resource Limited Settings”, describing critical steps and actions for effective implementation of a national PV policy: stakeholders’ engagement, statutory endorsement, institutionalization of the PV policy, addressing the roles and responsibilities of stakeholders and securing funding (52).

## DISCUSSION

This scoping review was the first, to the best of our knowledge, to describe the strategies and interventions to improve PV capacity in LMIC in the last three decades. Two-thirds of papers identified in this review described interventions aimed at improving ADR reporting. Educating HCP on PV and ADR reporting is an effective method to change their attitude towards reporting, and provides them with an understanding of the ADR reporting process and other issues associated with underreporting of ADRs, such as fear of litigation (53, 54). It is not surprising, therefore, given the major role played by HCP in reporting of ADRs, that a majority of interventions identified in this review had an educational component. Findings revealed that these PV educational or training workshops were very effective in improving PV knowledge with a corresponding increase in the number and quality of ADR reports. This is in line with numerous previous scoping or systematic reviews aimed at mapping interventions to improve ADR reporting among HCP (8, 9).

Several papers also described training of other PV stakeholders such as the national safety review committees (46, 55-57) and consumers (30, 37, 58, 59). National safety review committees perform causality assessments of serious cases and support analysis of national PV data. Nevertheless, the timing of the causality assessment is important and their work is dependent on stable funding. ADRs from patients and consumers are an important source of information on the safety and effectiveness of medicines and provides useful information for improving drug safety and health care from a different perspective (60). Consequently, there is a global trend toward educating and empowering patients and consumers to report ADRs, and studies have established the significant contribution of consumer reporting to signal detection (59, 61, 62).

Electronic reporting of ADRs is one of the main strategies used globally to improve spontaneous reporting of ADRs by HCP (63). In many developed countries, patients and consumers can directly report ADRs through web-based platforms. As seen in this review, NRA in LMIC are also moving towards implementing web-based electronic reporting and data management platforms such as Vigibase™ (34, 35) to handle data at the national level. Likewise, mobile phones and mobile phone applications are increasingly used to expand and facilitate ADR reporting in HCP as demonstrated by several papers in this review (32, 33, 36, 39, 64). Enhance passive and active surveillance are PV methods usually implemented during new medicinal production to increase and improve the collection and quality of PV data. The introduction of novel vaccines and drugs in LMIC represents an opportunity to strengthen national safety surveillance systems. Significant technical and financial efforts from national and global PV stakeholders are directed towards enhancing national PV systems prior to medicine introduction. In addition to other interventions, training of HCP and sometimes, patients, on PV and ADRs is one of the key preparatory activities of vaccine introduction preparations, often implemented through passive, enhanced passive and active surveillance as identified in this review (36, 37, 39).

Educating HCP and other PV actors on different aspects of PV and ADR reporting remains an essential aspect of building national PV capacity and numerous organisations and NRA provide diverse PV training programmes. These types of interventions are often simple to implement and can attain HCP at all levels of the health system. However, mechanisms for measuring their long-term impact are needed and there is a need for retraining and continuous education to sustain acquired knowledge. Several papers in this review showed that the increase in ADRs observed after the intervention periods was not sustained in subsequent months (18, 22, 39), and only few publications mentioned retraining of HCP (29), mentorship and supervisory visits (28, 45, 64) as part of the interventions. Therefore, education alone does not suffice and could have more impact in combinations with other techniques and implemented within the broader context of strengthening national PV systems.

About one-third of papers identified presented a more comprehensive structured approach to develop national PV systems. When interventions are implemented in a structured manner and are supported by validated tools such as the WHO GBT, different components of the PV system can be systematically strengthened, by formulating institutional development plans that address identified gaps, prioritize interventions and facilitate monitoring of progress and achievements (5). As shown in the Results, the stepwise implementation of PV in India, targeting the key indicators of the PV system (40-42), resulted in the creation of a performant PV system assessed at maturity level 4 for several functions such as clinical trial oversight, marketing authorization and vigilance. India contributes 3% ADRs to the global safety database, with a completeness score of 0.93 out of 1. Likewise, in Malawi, a step-wise approach targeting different PV components, with the collaboration of different technical agencies, enabled the creation of a national PV system, with guidelines, organisational structure, PV infrastructure and mechanisms for ADR reporting and other PV processes such as active surveillance (44, 56, 65).

Though several papers described different strengthening interventions at the national level with positive results, it was more difficult to identify national strategies with holistic operational plans in line with recent WHO recommendations, such as identified in Uganda (47), Tanzania (66) and Nigeria (48, 67). A recent publication has described the key strategies, outcomes, challenges and lessons learnt on building Eritrean national PV system. The key strategies for success included: i) establishing organisational capacity and capability for PV; ii) integrating PV into all PHP; iii) integrating PV into academia; iv) maximizing ADR reporting through massive training, recognition and feedback, appointing PV focal points, a combination of passive, stimulated passive and active surveillance, amongst others; and v) signal detection and assessment. As a result, Eritrea has been rated among the top reporting countries in Africa and the centre has detected 30 safety signals, and achieved maturity level 3 on the WHO GBT (68). Such strategic documents on strengthening PV systems provide valuable information to national and global PV initiatives and stakeholders on developing PV systems in LMIC. These strategies can be adapted to specific local contexts with feasible operational plans, providing a framework to build, track and continuously improve all components of the national PV system.

Overall, findings from this review revealed similar strategies and interventions aimed at building functional PV systems across several LMIC. The main strategies identified were: i) educating HCP and increasing patients, on PV and reporting of ADRs; ii) facilitating reporting of ADRs using different mechanisms such as mobile and electronic reporting, enhanced passive and active surveillance; iii) integrating PV in PHP and developing mechanisms for coordination and information exchange; iv) establishing and training national safety committees and developing capacity for data analysis; v) developing PV policies, guidelines and tools; vi) including PV in curriculum of HCP. There was a paucity of articles describing capacity building for more advanced PV activities such as establishing quality systems with procedures for aggregate data analysis and signal detection. Nevertheless, interventions identified, whether alone or in combination, resulted in either an increase in PV knowledge, ADR reporting or improvement of national PV systems or capacity, though not necessarily resulting in functional PV systems.

### Challenges in strengthening ADR reporting and PV systems

The main challenges identified in papers included in this review were:

1. Underreporting of ADRs, associated with numerous factors including lack of/or inadequate knowledge of reporting procedures, unavailability of reporting forms, lack of motivation, and lack of adequate methods of communication (29, 40, 58, 59, 69).
2. Lack of financial and human resources dedicated to PV, primarily because PV activities in numerous LMIC rely largely on external funds. When these funds run out, PV activities also stagnate (45, 46).
3. Insufficient national coordination of PV activities and inadequate collaboration and coordination between major national PV stakeholders hinders PV development (43, 44, 46).
4. Interventions were not sustained because often, there were no long-term plans and continuous funding. Only few publications mentioned retraining of HCP, mentorship and supervisory visits and these often come at a cost (22, 26, 29, 70).

### Lessons learnt and implications of the findings for research

As revealed in the current and in previous reviews, a majority of interventions to improve spontaneous reporting in LMIC were based on training workshops, often single-centre short-term activities. The importance of spontaneous reporting and educational interventions to improve passive surveillance cannot be over-emphasized. However, these should ideally be continuous, and combined with other activities aimed at developing national PV systems. Findings show a gap in more advanced PV activities such as aggregate data analysis, signal detection, active safety surveillance and risk management and communication. With novel medicines increasingly introduced in low-resource settings, effective and adequate safety surveillance will depend on these countries’ capacity for signal detection, analysis and interpretation of safety data. Therefore, building capacity for advanced PV activities is an important area of investment for future PV strengthening initiatives. Lastly, improving PV systems should ideally be holistic, targeting all components of the PV system, with plans for sustainability, and procedures to periodically assess improvements with validated tools.

### Limitations of the research

This review aimed to identify strategies and interventions to build and strengthen PV systems in LMIC. The search strategy mainly identified interventions aimed at improving ADR knowledge and reporting amongst HCP, with fewer articles identified on strategic approaches to strengthen national PV systems in LMIC. This is probably because results of such strategies and activities are not often published in scientific literature and are uncommonly publicly available. In addition, there is also a possible publication bias with the effect that only reasonably successful interventions at the national level would likely be submitted for publication. A limited search of selected national regulatory websites was conducted, because an extended search of NRA websites would have required significant resources, beyond the scope of this research, as discussed in Methods.

## CONCLUSIONS

Findings from this review showed that a combination of interventions such as education, financial incentives, mobile and electronic reporting, and enhanced passive and active surveillance significantly improved ADR reporting. Educating HCP on PV was the most common approach to build PV capacity in LMIC. However, education alone is insufficient in that it must be continuous, and ideally organised within the framework of strengthening the national PV system. A structured approach, targeting all PV components, with a particular focus on building capacity for more complex PV activities, is judicious and provides a framework to continuously build, track and improve national PV strengthening activities.

## Supporting information

Additional file with appendices

## Data Availability

All data produced in the present work are contained in the manuscript and in the additional file

## STATEMENTS AND DECLARATIONS

### Ethics approval

The Ethikkommission Nordwest-und Zentralschweiz (EKNZ) has confirmed that no ethical approval was required.

### Author’s contributions

**Olga Menang:** Conceptualization, data curation, formal analysis, methodology, project administration, resources, visualization, writing – original draft, writing – review & editing

**Christian Burri, Andrea Kuemmerle and Karen Maigetter** contributed to the study design and the analysis and interpretation of the data. All authors read and approved the final manuscript.

## ACKNOWLEDGEMENTS

The authors also wish to thank **Andy Stergachis** (Director, Global Medicines Program & Biomedical Regulatory Affairs Program, University of Washington School of Pharmacy) and **Günther Fink** (Head, Household Economics and Health System Research Unit, SwissTPH) for reviewing this manuscript. Thanks to **Hannah Ewald** (University Medical Library, University of Basel, Switzerland) for advice on the research protocol, developing the search strategy and for conducting the literature searches. Thanks to **Niranjan Bhat** (PATH) and **Emmanuel Mpinga Kabengele** (Institute of Global Health at the University of Geneva) for reviewing the scoping review.

