## Additional file with appendices for "Strategies and interventions to strengthen pharmacovigilance systems in low- and middle-income countries: a scoping review"

**ADDITIONAL FILES**

Authors

Olga Menang^1,2,^ , Andrea Kuemmerle^1,2,^ , Karen Maigetter ^1,2,^ Christian Burri^1,2^

^1^Swiss Tropical and Public Health Institute, Department of Medicine, Allschwil, Switzerland;

^2^University of Basel, Basel, Switzerland;

Corresponding to Olga Menang (ORCID ID 0000-0002-3928-4288):

### **Appendix I: Search strategy in Ovid MEDLINE**

((afghanistan OR albania OR algeria OR american samoa OR angola OR "antigua and barbuda" OR antigua OR barbuda OR argentina OR armenia OR armenian OR aruba OR azerbaijan OR bahrain OR bangladesh OR barbados OR republic of belarus OR belarus OR byelarus OR belorussia OR byelorussian OR belize OR british honduras OR benin OR dahomey OR bhutan OR bolivia OR "bosnia and herzegovina" OR bosnia OR herzegovina OR botswana OR bechuanaland OR brazil OR brasil OR bulgaria OR burkina faso OR burkina fasso OR upper volta OR burundi OR urundi OR cabo verde OR cape verde OR cambodia OR kampuchea OR khmer republic OR cameroon OR cameron OR cameroun OR central african republic OR ubangi shari OR chad OR chile OR china OR colombia OR comoros OR comoro islands OR iles comores OR mayotte OR democratic republic of the congo OR democratic republic congo OR congo OR zaire OR costa rica OR "cote d’ivoire" OR "cote d’ ivoire" OR cote divoire OR cote d ivoire OR ivory coast OR croatia OR cuba OR cyprus OR czech republic OR czechoslovakia OR djibouti OR french somaliland OR dominica OR dominican republic OR ecuador OR egypt OR united arab republic OR el salvador OR equatorial guinea OR spanish guinea OR eritrea OR estonia OR eswatini OR swaziland OR ethiopia OR fiji OR gabon OR gabonese republic OR gambia OR "georgia (republic)" OR georgian OR ghana OR gold coast OR gibraltar OR greece OR grenada OR guam OR guatemala OR guinea OR guinea bissau OR guyana OR british guiana OR haiti OR hispaniola OR honduras OR hungary OR india OR indonesia OR timor OR iran OR iraq OR isle of man OR jamaica OR jordan OR kazakhstan OR kazakh OR kenya OR "democratic people’s republic of korea" OR republic of korea OR north korea OR south korea OR korea OR kosovo OR kyrgyzstan OR kirghizia OR kirgizstan OR kyrgyz republic OR kirghiz OR laos OR lao pdr OR "lao people's democratic republic" OR latvia OR lebanon OR lebanese republic OR lesotho OR basutoland OR liberia OR libya OR libyan arab jamahiriya OR lithuania OR macau OR macao OR republic of north macedonia OR macedonia OR madagascar OR malagasy republic OR malawi OR nyasaland OR malaysia OR malay federation OR malaya federation OR maldives OR indian ocean islands OR indian ocean OR mali OR malta OR micronesia OR federated states of micronesia OR kiribati OR marshall islands OR nauru OR northern mariana islands OR palau OR tuvalu OR mauritania OR mauritius OR mexico OR moldova OR moldovian OR mongolia OR montenegro OR morocco OR ifni OR mozambique OR portuguese east africa OR myanmar OR burma OR namibia OR nepal OR netherlands antilles OR nicaragua OR niger OR nigeria OR oman OR muscat OR pakistan OR panama OR papua new guinea OR new guinea OR paraguay OR peru OR philippines OR philipines OR phillipines OR phillippines OR poland OR "polish people's republic" OR portugal OR portuguese republic OR puerto rico OR romania OR russia OR russian federation OR ussr OR soviet union OR union of soviet socialist republics OR rwanda OR ruanda OR samoa OR pacific islands OR polynesia OR samoan islands OR navigator island OR navigator islands OR "sao tome and principe" OR saudi arabia OR senegal OR serbia OR seychelles OR sierra leone OR slovakia OR slovak republic OR slovenia OR melanesia OR solomon island OR solomon islands OR norfolk island OR norfolk islands OR somalia OR south africa OR south sudan OR sri lanka OR ceylon OR "saint kitts and nevis" OR "st. kitts and nevis" OR saint lucia OR "st. lucia" OR "saint vincent and the grenadines" OR saint vincent OR "st. vincent" OR grenadines OR sudan OR suriname OR surinam OR dutch guiana OR netherlands guiana OR syria OR syrian arab republic OR tajikistan OR tadjikistan OR tadzhikistan OR tadzhik OR tanzania OR tanganyika OR thailand OR siam OR timor leste OR east timor OR togo OR togolese republic OR tonga OR "trinidad and tobago" OR trinidad OR tobago OR tunisia OR turkey OR turkmenistan OR turkmen OR uganda OR ukraine OR uruguay OR uzbekistan OR uzbek OR vanuatu OR new hebrides OR venezuela OR vietnam OR viet nam OR middle east OR west bank OR gaza OR palestine OR yemen OR yugoslavia OR zambia OR zimbabwe OR northern rhodesia OR global south OR africa south of the sahara OR sub-saharan africa OR subsaharan africa OR africa, central OR central africa OR africa, northern OR north africa OR northern africa OR magreb OR maghrib OR sahara OR africa, southern OR southern africa OR africa, eastern OR east africa OR eastern africa OR africa, western OR west africa OR western africa OR west indies OR indian ocean islands OR caribbean OR central america OR latin america OR "south and central america" OR south america OR asia, central OR central asia OR asia, northern OR north asia OR northern asia OR asia, southeastern OR southeastern asia OR south eastern asia OR southeast asia OR south east asia OR asia, western OR western asia OR europe, eastern OR east europe OR eastern europe OR developing country OR developing countries OR developing nation? OR developing population? OR developing world OR less developed countr* OR less developed nation? OR less developed population? OR less developed world OR lesser developed countr* OR lesser developed nation? OR lesser developed population? OR lesser developed world OR under developed countr* OR under developed nation? OR under developed population? OR under developed world OR underdeveloped countr* OR underdeveloped nation? OR underdeveloped population? OR underdeveloped world OR middle income countr* OR middle income nation? OR middle income population? OR low income countr* OR low income nation? OR low income population? OR lower income countr* OR lower income nation? OR lower income population? OR underserved countr* OR underserved nation? OR underserved population? OR underserved world OR under served countr* OR under served nation? OR under served population? OR under served world OR deprived countr* OR deprived nation? OR deprived population? OR deprived world OR poor countr* OR poor nation? OR poor population? OR poor world OR poorer countr* OR poorer nation? OR poorer population? OR poorer world OR developing econom* OR less developed econom* OR lesser developed econom* OR under developed econom* OR underdeveloped econom* OR middle income econom* OR low income econom* OR lower income econom* OR low gdp OR low gnp OR low gross domestic OR low gross national OR lower gdp OR lower gnp OR lower gross domestic OR lower gross national OR lmic OR lmics OR third world OR lami countr* OR transitional countr* OR emerging economies OR emerging nation?).ti,ab,sh,kf. OR (afghan OR afghans OR afghani OR albanian? algerian? OR american samoan? OR angolan? OR antiguan? OR barbudan? OR argentine? OR argentinian? OR argentinean? OR armenian? OR aruban? OR azerbaijani? OR bahraini? OR bangladeshi? OR bangalees OR bajan? OR belarusian? OR byelorussian? OR belizean? OR beninese? OR bhutanese OR bolivian? OR bosnian? OR botswana OR batswana OR brazilian? OR brasilian? OR bulgarian? OR burkinabe OR burkinese OR burundian? OR cape verdean? OR cabo verdean? OR cambodian? OR khmer OR cameroonian? OR central african? OR chadian? OR chilean? OR chinese OR colombian? OR comorian? OR congolese OR costa rican? OR ivorian? OR croatian? OR cuban? OR cypriot? OR czech? OR djiboutian? OR dominican? OR ecuadorian? OR egyptian? OR salvadoran? OR equatorial guinean? OR equatoguinean? OR eritrean? OR estonian? OR swazi? OR swati? OR ethiopian? OR fijian OR gabonese OR gabonaise OR gambian? OR georgian? OR ghanaian? OR gibraltarian? OR greek? OR grenadian? OR guamanian? OR guatemalan? OR guinean? OR bissau guinean? OR guyanese OR haitian? OR honduran? OR hungarian? OR indian? OR indonesian? OR iranian? OR iraqian? OR iraqi? OR manx OR jamaican? OR jordanian? OR kazakhstani? OR kenyan? OR kirabati OR kirabatian? OR north korean? OR korean? OR kosovar? OR kosovan? OR kyrgyz* OR lao OR laotian? OR latvian? OR lebanese OR lesothan? OR lesothonian? OR mosotho OR basotho OR liberian? OR libyan? OR lithuanian? OR macanese OR macedonian? OR malagasy OR madagascan? OR malawian? OR malaysian? OR maldivian? OR malian? OR maltese OR marshallese? OR mauritanian? OR mauritian? OR mexican? OR micronesian? OR moldovan? OR mongolian? OR mongol OR montenegrin? OR moroccan? OR mozambican? OR burmese OR myanma OR namibian? OR nauruan? OR nepali OR nepalese OR netherlands antillean? OR nicaraguan? OR nigerien? OR nigerian? OR northern mariana islander? OR mariana? OR omani? OR pakistani? OR palauan? OR panamanian? OR papua new guinean? OR paraguayan? OR peruvian? OR philippine? OR philipine? OR phillipine? OR phillippine? OR filipino? OR filipina? OR polish OR pole OR poles OR portuguese OR puerto rican? OR romanian? OR russian? OR soviet people OR soviet population OR rwandan? OR rwandese OR ruandan? OR ruandese OR samoan? OR sao tomean? OR santomean? OR saudi arabian? OR saudi? OR senegalese OR serbian? OR montenegrin? OR seychellois OR seychelloise? OR sierra leonean? OR slovak? OR slovene? OR solomon islander? OR somali? OR south african? OR south sudanese OR sri lankan? OR ceylonese OR kittitian? OR nevisian? OR saint lucian? OR vincentian? OR sudanese OR surinamese? OR syrian? OR tajik? OR tajikistani? OR tanzanian? OR tanganyikan? OR thai OR timorese? OR togolese OR tongan? OR trinidadian? OR tobagonian? OR tunisian? OR turk? OR turkish OR turkmen? OR tuvaluan? OR ugandan? OR ukrainian? OR uruguayan? OR uzbek? OR vanuatu* OR venezuelan? OR vietnamese OR yemeni? OR yemenite? OR yemenese OR yugoslav? OR yugoslavian? OR zambian? OR zimbabwean?).ti,ab,sh,kf.)

AND (((exp adverse drug reaction reporting systems/ OR exp pharmacovigilance/ OR exp product surveillance, postmarketing/ OR ((exp drug-related side effects and adverse reactions/ OR exp adverse effects/ OR exp adverse drug event/) AND (report*.ti,ab. OR system.ti,ab. OR systems*.ti,ab. OR monitor*.ti,ab. OR surveill*.ti,ab.))) AND (increas*.ti,ab. OR improv*.ti,ab. OR enhanc*.ti,ab. OR promot*.ti,ab. OR education.ti,ab. OR strengthen*.ti,ab. OR capacity building.ti,ab. OR building capacity.ti,ab. OR implement*.ti,ab. OR operat*.ti,ab. OR develop*.ti,ab. OR building*.ti,ab.)) OR ((pharmacovigilance.ti,ab. OR adverse drug reaction reporting systems.ti,ab. OR pharmacovigilance program*.ti,ab. OR pharmacovigilance activit*.ti,ab. OR adverse drug event surveillance system.ti,ab. OR adverse event monitoring.ti,ab. OR vaccine safety system*.ti,ab. OR drug safety.ti,ab. OR vaccine safety.ti,ab. OR safety surveillance .ti,ab. OR pharmacogovernance.ti,ab. OR ((adverse event*.ti,ab. OR adverse drug reaction*.ti,ab. OR side effect*.ti,ab. OR adverse effect*.ti,ab. OR adverse reaction*.ti,ab. OR undesirable event*.ti,ab. OR adverse event following immunization.ti,ab.) ADJ2 (report*.ti,ab. OR system.ti,ab. OR systems*.ti,ab. OR monitor*.ti,ab. OR surveill*.ti,ab.))) ADJ3 (increas*.ti,ab. OR improv*.ti,ab. OR enhanc*.ti,ab. OR promot*.ti,ab. OR education.ti,ab. OR strengthen*.ti,ab. OR capacity building.ti,ab. OR building capacity.ti,ab. OR implement*.ti,ab. OR operat*.ti,ab. OR develop*.ti,ab. OR building*.ti,ab.)))

### **Appendix II: Sources of grey literature search**

*Search terms use: pharmacovigilance strategy, pharmacovigilance strengthening*

| **Source** | **Organisation** | **Website** |
| --- | --- | --- |
| Brazil | Anvisa | <https://www.gov.br/anvisa/pt-br> |
| Burkina Faso | Direction Générale de la Pharmacie, du Médicament et des Laboratoires Direction | <https://www.servicepublic.gov.bf/contact/direction-generale-de-la-pharmacie-du-medicament-et-des-laboratoires> |
| Cameroon | Direction de la Pharmacie du Médicament et des Laboratoires (DPML) | [https://dpml.cm/index.php/en/home-en#](https://dpml.cm/index.php/en/home-en) |
| Chile | ISP | <https://www.ispch.gob.cl/> |
| Côte d'Ivoire | Autorité Ivoirienne de Régulation Pharmaceutique | <https://www.airp.ci/frGahana> |
| Ghana | Food and Drugs Authority | <http://www.fdaghana.gov.gh/> |
| India | Central Drugs Standard Control Organization | <https://cdsco.gov.in/opencms/opencms/en/PvPI/> |
| Kazakhstan | Ministry of Healthcare of the Republic of Kazakhstan | <https://www.gov.kz/memleket/entities/dsm?lang=en> |
| Kenya | Pharmacy and Poisons Board | <https://web.pharmacyboardkenya.org/pharmacovigilance/mala> |
| Malawi | Pharmacy and Medicines Regulatory Authority (PMRA) | <https://pmra.mw/> |
| Morocco | Centre Anti Poison et PV du Maroc | <http://www.capm-sante.ma/pv-pharmacovigilance> |
| Namibia | Namibia Medicines Regulatory Council | <https://nmrc.gov.na/> |
| Nigeria | National Agency for Food and Drug Administration and Control | <https://www.nafdac.gov.ng/> |
| Republic of Congo | Direction de la Pharmacie et du Médicament | <https://www.dpm-congo.net/> |
| Senegal | La Direction de la Pharmacie et du Médicament | <https://www.sante.gouv.sn/les-directions/la-direction-de-la-pharmacie-et-du-m%C3%A9dicament> |
| Serbia | Medicines and Medical Devices Agency of Serbia (ALIMS) | <https://www.alims.gov.rs/english/pharmacovigilance/> |
| South Africa | South African Health Products Regulatory Authority (SAHPRA)Uganda FDA | <https://www.sahpra.org.za/> |
| Tanzania | Tanzania Medicines and Medical Devices Authority (TMDA) | <https://www.tmda.go.tz/> |
| Uganda | National Drug Authority | <https://www.nda.or.ug/> |
|  | Uppsala Monitoring Centre (UMC | <https://who-umc.org/> |
|  | East African Community | <https://www.eac.int/medicines-regulatory-guidelines> |
|  | Tropical Disease Research | <https://tdr.who.int/> |
|  | PharmacoVigilance Africa (PAVIA) | <https://pavia-africa.net/> |
|  | World Health Organisation | <http://www.who.int/> |
|  | Systems for Improved Access to Pharmaceuticals and Services (SIAPS) Program | <https://siapsprogram.org/> |
|  | WHO Regional Office for Africa | <https://www.afro.who.int/> |
|  | WHO Regional Office for the Eastern Mediterranean | <http://www.emro.who.int/index.asp> |
|  | WHO Regional Office for Southeast Asia | <https://www.who.int/southeastasia> |
|  | Management Sciences for Health (MSH) | <https://msh.org/> |
|  | USAID | <https://www.usaid.gov/> |
|  | Bill and Melinda Gates Foundation | <https://www.gatesfoundation.org/> |
|  | The Centers for Disease Control and Prevention (CDC) | <https://www.cdc.gov/> |
|  | African Collaborating Centre  For Pharmacovigilance  & Surveillance (ACC) | <https://www.acc-afro.org/> |

### **Appendix III: Data extraction instrument**

| Author/ Country | Nature of intervention | Context/  setting | Description of improvement intervention | Results /  Outcomes | Challenges encountered | Lessons learnt and recommendations |
| --- | --- | --- | --- | --- | --- | --- |

### **Appendix IV: Context and setting**

| Region^1^ | Number of Publications | Countries |
| --- | --- | --- |
| East Asia Pacific region | 9 | China (3), Philippines (2),Malaysia (2),Vietnam (2) |
| Europe and Central Asia | 1 | Kazakhstan (1) |
| Latin America and the Caribbean | 6 | Bolivia (1),Brazil (2), Mexico (2), Multi-country (1) (Argentina, Chile, Colombia, Costa Rica, Honduras, Peru, and Uruguay) |
| Middle East and North Africa | 5 | Egypt (1), Iran (1), Jordan (1), Lebanon (1), Morocco (1) |
| South Asia | 17 | India (12), Nepal (3), Multi-country (2) :   - Bangladesh, Bhutan, India, Indonesia, Maldives, Nepal, Sri Lanka, Thailand, Timor-Leste and Vietnam - Bangladesh, Bhutan, DRP Korea, India, Indonesia, Maldives, Myanmar, Nepal, Sri Lanka, Thailand and Timor Este |
| Sub-Saharan Africa | 28 | Burkina Faso (1), Cameroon (2), Democratic Republic of the Congo (DRC) (1), Ethiopia (1), Malawi (2), Mozambique (2), Namibia (1), Nigeria (4), Rwanda (1), Senegal (1), South Africa (4), Sudan (1), Swaziland (2),Tanzania (2), Uganda (1) Multi-country (2):   - Ghana, Kenya and Malawi - Benin, Burkina Faso, Cameroon, Chad, Ethiopia, Ghana, Niger, Nigeria, Senegal, Sudan, and The Gambia |
| Global/LMIC | 4 | Global recommendations |
| Total | **70** |  |

^1^ According to World Bank region classification (1)

### **Appendix V: Nature of interventions and outcomes**

| Author country | Nature of intervention | Objective | Description of intervention | Outcome | Challenges encountered | Lessons learnt and recommendations |
| --- | --- | --- | --- | --- | --- | --- |
| 1. Interventions aimed at increasing PV knowledge and AE or ADR reporting | | | | | | |
| Abu Farha et al.,  Jordan  (2) | Educational | To evaluate the impact of an educational workshop on the knowledge and perception of HCP towards PV in a Jordanian tertiary teaching hospital. | - 1h educational workshop on PV | - Improvement in knowledge and perception scores | - The influence on the practice of ADRs reporting was not studied | - Continuous training and different strategies needed to encourage adherence to appropriate PV practices |
| Agu et al.,  Nigeria  (3) | Mixed | To evaluate the change in knowledge, attitudes and practices of healthcare professionals about ADR monitoring and reporting after six months of capacity building interventions in a Nigerian tertiary hospital. | - Five–day group training on PV for antiretroviral (ARV) drugs - Establishing a multidisciplinary ARV PV Committee to coordinate PV of ARV in the hospital - Regular feedback and data review - Dissemination of PV standard operating procedures and reporting forms | - Increase in mean knowledge and attitude scores at post-intervention - Increase in number of HCP reporting ADRs - 163 yellow cards were received from 49 pharmacists | - Lack/inadequate knowledge on ADR reporting - Unavailability of reporting forms - Ignorance of reporting procedures | - HCP were more willing to practice PV following the intervention |
| Albadawi et al., Sudan  (4) | Educational | To evaluate HCP’s knowledge and attitude towards PV among health professionals in Ribat University Hospital, Sudan and to assess the impact of an intervention. | - PV lecture sessions - Pamphlets - Mobile phones reminders or posters | - Mean PV knowledge and attitude significantly improved post-intervention | None reported | None reported |
| Alraie et al.,  Egypt  (5) | Educational | To assess the impact of PV awareness workshop on knowledge of hospital pharmacists; and to identify the main factors and barriers that influence ADR reporting. | - Three-day PV awareness workshop - A follow-up phone call after three months to detect barriers encountered - Monitoring and analysis of ADR reports for six months | - Increased knowledge and awareness after intervention | - Reporting hindered by lack of time and administrative barriers - Inability to complete patient details | - Limited number ADR reports in the 6 months follow-up period, indicating that education alone does not suffice to promote reporting |
| Ateudjieu et al.,  Cameroon  (6) | Mixed | To assess the effect of weekly SMS and weekly supervisory visits on AEFI reporting rate during a meningitis immunization campaign conducted in Cameroon in 2012 using the meningitis A conjugate vaccine (MenAfriVacTM). | - A weekly SMS reminder to actively detect and report AEFI - Weekly supervision by a nurse to support with AEFI detection and reporting - No intervention | - The incidence of AEFI reported per 100 health facilities per week was 20.0 in the SMS group, 40.2 in supervision group and 13.6 in the control group. - Supervision led to a significant increase of AEFI reporting rate compared to SMS and control group | - Low completion rate of AEFI reporting forms | - Supervision should be included as an intrinsic part of planning AEFI surveillance during vaccination campaigns |
| Bepari et al.,  India  (7) | Educational | To evaluate the impact of a multi-faceted educational intervention on the knowledge, perception, and practice skills of PV among undergraduate pharmacy students. | - Lecture on PV concepts with practical lessons on completing ADR forms - Distribution of educational material to participants | - Significant improvement in the knowledge, perceptive behaviour, and practice skills scores of PV | None reported | None reported |
| Bisht et al.,  India  (8) | Educational | To assess the level of knowledge, attitude, and the practices of PV among doctors attending educational training for improving awareness of PV. | - A lecture to increase awareness of ongoing PV program in the institute. | - Significant increase in the knowledge and awareness of PV though no major difference in practice | None reported | - Continuous education and others strategies needed to improve the awareness and reporting of ADRs |
| Bravo  Alcantero et al.,  Argentina, Chile, Colombia, Costa Rica, Honduras, Peru, and Uruguay  (9) | Passive and active safety surveillance systems | To build capacity for active surveillance of vaccine adverse events in the Americas based on the evaluation of two well-established relationships, the risk of immune thrombocytopenic purpura and aseptic meningitis following first dose of measles, mumps and rubella vaccines. | - Distance learning exercise to test data collection and quality - Retrospective standardized data collection of all adverse events of special interest (AESIs) over a four-year period | - The study contributed 79 AESIs to the WHO international proof-of-concept hospital-based active surveillance system | - Significant lag times between activities - Lack of access to vaccination records for all confirmed cases in a few sentinel sites | - Active safety surveillance systems may be sustainable if they are integrated into national health systems |
| Chang et al.,  China  (10) | Mixed | Assess the effectiveness of a financial intervention based on a fine and a bonus for improving spontaneous reporting of ADRs by physicians in a hospital setting. | - Pre intervention: implantation of the ADR database - First intervention: financial incentive - Second intervention: financial incentive and strict regulations for antimicrobial agents | - Significant increase in ADRs (29 at pre-intervention; 277 at first intervention; and 666 at second intervention period) - Increase in the quality of reports | None reported | None reported |
| Cortes Serra et al.,  Bolivia  (11) | Educational | To strengthen the Bolivian PV system, focusing on Chagas disease and Tuberculosis. | - Design and implementation of new reporting form - Specific training on drug safety monitoring and ADR reporting - Three follow-up visits - Evaluation of the effectiveness of interventions | - No differences in reporting rates between the reporting form developed by the NRA and the new form | - Lack of knowledge on PV and ADR reporting - Insufficient training - HCP possibly did not see the benefits of using the new tool - Lack of internet access - Long-term effect cannot be evaluated yet | - Reinforcement of the Bolivian PV system should involve a long-term perspective and the engagement of national stakeholders at all levels |
| Damdar et al.,  (12) | Educational | To analyse the impact of educational intervention on the knowledge of PV. | - A workshop on filling the spontaneous ADR reporting form and demonstration of WHO and Naranjo causality assessment scale | - Increase in knowledge and awareness of PV monitoring and reporting ADRs | None reported | None reported |
| Deepalakshmi et al.,  India  (13) | Educational | To educate and train the community pharmacists on PV and assess the knowledge, attitude and practice of ADR monitoring and implementation of ADR reporting in their practice. | - Distribution and filling of ADR forms - PV awareness trainings - ADR awareness posters displayed at community pharmacies - Follow-up after 6 months to assess the changes if any changes in practice | - Implementation of ADR monitoring services in selected community pharmacies - Significant increase in knowledge and practice of PV | None reported | None reported |
| Elkalmi et al.,  Malaysia  (14) | Educational | To assess the knowledge and perception of community pharmacists in Malaysia towards the reporting of ADRs and to evaluate the effectiveness of an educational program to improve knowledge on ADR reporting. | - Implementation of an educational program on PV and ADR reporting for community pharmacists | - Significant increase in mean PV knowledge scores and ADR reporting compared to the baseline | Low participation rate | None reported |
| Fang et al.,  China  (15) | Mixed | To investigate changes in spontaneous reporting compliance and ADR patterns following adoption of a new hospital reporting system, and multiple interventions designed for its improvement use under modified drug administration guidelines. | - Implementation of new guidelines on antibiotic use and ADR reporting - Financial incentives to the reporting physician and department - Training on PV and ADR reporting - Improvement of the computer system - Regular publishing of ADR information | - No significant difference in reporting rates pre- and post- intervention - Significant increase in total reports - Improvements in reporting compliance - Increase in reporting of serious ADRs | None reported | None reported |
| Gebreyohannes et al.,  Ethiopia  (16) | Educational | Assess whether pictorial intervention would help to identify and improve ADR reporting in an ARV clinic in Northwest Ethiopia | - Evaluation of a pictogram-enhanced tool to identify and report ADRs. | - Pictorial representation resulted in only slight improvement in identification and reporting of ADRs | The study was conducted in a single centre, and most of the participants were from low to middle socioeconomic class, | - Pictograms can be used to educate about ADRs associated with ARV |
| He et al.,  China  (17) | Mixed | To increase the number of ADR reports and promote hospital PV through a pharmacists-led ADR management model. | - Strengthening information and education in key departments - Establishing an internal hospital network direct reporting system | - Significant increase in the quantity (177 pre-intervention vs 533 post-intervention) and quality of ADRs reported | The model has high requirements for the professional ability of pharmacists. | None reported |
| Ithnin et al.,  Malaysia  (18) | Mobile and electronic reporting | To develop and assess utility of mobile apps in assisting clinical decision in ADR assessments of causality, severity, and preventability using validated tools. | - Development of ADR causality assessment App - Testing and measuring the App's usability | - Developed apps made available in Google Play Store - 609 users across different countries downloaded the App - Users assessed the App's usability as acceptable | None reported | None reported |
| Jha et al.,  Nepal  (19) | Educational | To examine community pharmacists' knowledge and attitude about pharmacovigilance before and after an educational intervention. | - 2h Training on PV - Poster sessions | - Improvement in knowledge and attitude towards PV | - HCP lack time for ADR reporting - Lack of continuous feedback and information sharing on ADRs | None reported |
| Kabanywanyi et al.,  Tanzania  (20) | Passive and active safety surveillance systems | To identify and implement strategies that help meet safety monitoring requirements in observational study for artemether-lumefantrine administered as first-line treatment for uncomplicated malaria in rural Tanzania. | - Implementation of passive and active safety surveillance systems - SMS reporting - Training of HCP | - Increase in AEs reported | None reported | - SMS could provide a solution to communication challenges |
| Kadima et al.,  Rwanda  (21) | Mobile and electronic reporting | To explore the feasibility of outpatients to self-report ADRs using mobile phone technology and to picture such ADRs and medications received. | - 80 patients educated to report ADRs using mobile phones either by accepting a call from the investigators or calling directly | - Compared to the previous months where no ADRs were reported, 34 patients reported an ADR | None reported | - The use of a mobile phone could complement reporting of ADRs and enhance PV |
| Khalili v  Iran  (22) | Educational | Evaluation of clinical pharmacists’ interventions in improvement of knowledge, attitude and perception about ADRs in a teaching hospital. | - Training workshops 3 hour weekly for four weeks - Continuous sensitization on ADRs every other day for 1 month - Practice on completing yellow card | - Improvement in recognition and reporting of ADRs. | - Unawareness of the existence of a national ADR reporting system - Under-reporting due to insufficient information | - Health systems must have PV training and guidance on how to report ADRs |
| MacDonald et al.,  Bangladesh, Bhutan, India, Indonesia, Maldives, Nepal, Sri Lanka, Thailand, Timor-Leste  Vietnam  (23) | Educational | Enhance regional capacity to evaluate investigated AEFI and carry out causality assessment of serious AEFI previously assessed by country committees. | - Three-day 10 country workshop to strengthen causality assessment - Group work to review and assess cases AEFI - Interactive discussions on to improve AEFI investigation and causality assessment in the region | - Needs identification and improvement in AEFI investigations and causality. | None reported | - LMICs need WHO AEFI tools adapted to better fit LMIC - Importance of data sharing and networking |
| MacDonald et al.,  Bangladesh, Bhutan, DRP Korea, India, Indonesia, Maldives, Myanmar, Nepal, Sri Lanka, Thailand and Timor Este  (24) | Educational | To strengthen regional and in-country vaccine safety capacity, with a strong focus on related communication activities. | - Three-day workshop to strengthen causality assessment - Group work to review and assess cases AEFI - Interactive discussions on to improve AEFI investigation and causality assessment in the region | - Compared to 2014, AEFI detection and causality assessment skills had improved | None reported | - Numerous recommendations for SEAR countries, WHO Southeast Asia Regional Office, and for Global Partners |
| Mehta et al.,  South Africa  (25) | Passive and active safety surveillance systems | Implementation of a PV strategy to pilot locally relevant surveillance methods for detecting serious ADRs and signals related to artesunate+sulfadoxine/pyrimethamine. | - Home follow-up of patients by malaria control staff - Enhanced spontaneous reporting of ADRs - Active hospital surveillance for malaria-related admissions - A confidential enquiry into malaria-related deaths - Adverse events monitoring during two therapeutic efficacy studies | - Identification of only very few fatal cases and ADRs through home visits, enhanced spontaneous reporting, efficacy study, and active surveillance | - ADRs not reported to the malaria control programme were not identified - Inadequate follow-up - Insufficient diagnosis capacity - Inadequacy of medical records - Recall bias among the next of kin of deceased individuals | - A more integrated approach to disease surveillance that could incorporate elements of PV, resistance monitoring and rumour surveillance |
| Morales Rio et al.,  Mexico  (26) | Educational | Assess the effectiveness of a comprehensive intervention coordinated by a pharmacist. | - 1h group informative talk on PV - Reminders to medical staff during medical visits or retrospective review of medical records - Feedback to medical staff Improve accessibility to the ADR reporting forms | - Seven-fold increase in ADR identification after intervention - Fourteen-fold increase in ADR reporting compared to pre-intervention - The effect on both variables was maintained 6 months after the intervention | - Medical records review was expensive – time and human resource-consuming - Underreporting of ADRs | - Pharmacists play an important role in ADR reporting |
| MSH  Mozambique  (27) | Mobile and electronic reporting | To implement PViMS, a web-based PV tool for monitoring medicine safety. | - Three-day training on active surveillance of patients on the new dolutegravir-based ARV medicine. - Implementation of active surveillance and spontaneous reporting using PViMS | None reported | None reported | None reported |
| Ndiaye et al.,  Senegal  (28) | Passive and active safety surveillance systems | This study aimed to determine if AE reporting could be improved using a smartphone application provided to community health workers (CHW), or by active follow-up using a symptom card provided to caregivers. | - Enhanced spontaneous reporting via daily SMS reminders to HCP - Active surveillance through home visits to detect and collect AEs - Training of facility staff and CHWs on PV and reporting of ADR via smartphone application - Community sensitizations - Training on causality assessment - Operationalization of a safety database | - Increased incidence with enhanced reporting, and active surveillance - Improved timeliness of notifications - Despite increased surveillance, no serious adverse drug reactions were detected | - The incidence of AEs decreased in successive rounds of Seasonal Malaria Chemoprevention | - Involving CHWs in safety reporting, and training nurses and CHWs to report using a mobile phone application, can be used to enhance safety reporting and improve timeliness of notifications |
| Nkonde et al.,  South Africa  (29) | Educational Procedural | To implement interventions to strengthen the PV surveillance system for ADR reporting at Dr. George Mukhari Academic Hospital. | - Retrospective quantitative and qualitative review of ADR reports 9 months prior to interventions - Supportive training sessions for HCP on PV and ADR reporting - Posters about ADRs and ADR reporting procedures in hospital - Feedback on ADR reporting to HCP through newsletters and meeting - Review of ADR reports for quality and completeness | - Significant increase in number (one compared to 14 after the interventions) and completeness of ADRs reported. | - Low uptake of the implemented interventions due to lack of support from hospital management - Damage to and misuse of ADR boxes placed in the wards to facilitate ADR reporting - Poor attitude toward PV and resistance to change routines | - Training interventions should be continuous - A coordinated approach with different stakeholders would yield better results. - PV training should be incorporated into HCP undergraduate programmes - Introducing SOPs on ADR reporting is essential |
| Osakwe et al.,  Nigeria  (30) | Educational | To determine the knowledge and practice of PV amongst HCP in Nigeria and the impact of previous training in PV on their knowledge and practice. | - Email distribution of quarterly PV newsletters - Formal face-to face training | - Respondent’s knowledge and practice of PV was generally inadequate despite the fact that about a third were trained on PV | None reported | - Innovations in PV training methods are necessary for better impact |
| Poirot et al.,  Swaziland  (31) | Mixed | To assess the feasibility and acceptability of using a simple PV tool to collect standardized safety data in persons prescribed primaquine for the treatment of P. falciparum malaria. | - A standardized form to support the surveillance of possible adverse events following primaquine treatment - A patient information card to enhance awareness of known adverse drug reactions of primaquine - A database compiling recorded information, was developed and piloted. | - The study promoted and facilitated the capture of treatment emergent AEs after initiation of primaquine | - Underreporting was higher than anticipated - No baseline assessment of symptoms in order to reduce the workload for healthcare providers - Recall bias | None reported |
| Prakash et al.,  India  (32) | Mobile and electronic reporting | To develop an indigenous Google based Android mobile application known as “ADR PvPI” and to analyse the ADR related data reported through this mobile application on pilot basis. | - Implementation an android based mobile app for ADR reporting | - 5500 users have downloaded the app with an average rating of 4.26 - Significant increase in frequency of ADR reporting over the past year" | None reported | None reported |
| Rouamba et al.,  Burkina Faso  (33) | Passive and active safety surveillance systems | To monitor the occurrence of AEs after the use of artemisinin-based combination therapies (ACTs), identify potential drivers of reporting suspected ADRs and monitor AEs among women who were inadvertently exposed to ACTs in the first trimester of pregnancy. | - Passive safety surveillance - •Active safety surveillance - Community sensitization | - The Health and Demographic Surveillance System (HDSS) platform was useful in enabling passive and active surveillance of AEs | - Recall bias for patient reporting - Frequent stock outs of ACT drugs - Estimation of the incidence of AEs did not systematically take into consideration anomalies in biological parameters | - Active surveillance through an HDSS platform seems to be an achievable approach to collect safety data in rural areas |
| Sanghavi et al.,  India  (34) | Educational | To evaluate the baseline knowledge, attitude and practice of the doctors on ADR monitoring and PV, to identify reasons for under reporting and plan an effective intervention to inculcate ADR reporting in clinical practice. | - 45 minutes lecture on PV | - Intervention improved the knowledge, attitude and awareness of the ADR reporting system. | - Under reporting due to lack of time and poor understanding of ADR reporting system | - Training could be improved with supportive tools such as job aids |
| Santosh et al.,  Nepal  (35) | Mixed | To provide an overview of PV and HCP perspectives on ADR reporting in Nepal and to provide recommendations on possible ways to improve ADR reporting. | - Training of HCP to increase awareness and encourage reporting - Design appropriate training tools to enhance knowledge of ADR reporting among HCP - Expand the current ADR monitoring program to cover the whole country - Establish a functional PV advisory committee to take any technical decision related to ADR monitoring and ADR reports and also to establish communication strategy - Encourage universities and academic institutions to provide training on PV | None reported | None reported | None reported |
| Sevene et al.,  Mozambique  (36) | Mixed | To examine the impact of training and monitoring of HCP, making supervisory visits and the availability of telecommunication and transport facilities on the implementation of a PV system. | - Training workshops on PV - Implementation of a ‘yellow card’ system for spontaneous reporting of ADRs - District focal person designated to facilitate communication between the health staff and the PV unit - Retraining after 5 months to reinforce learnings from the first workshop | - Fourteen months after the first training, 67 ADR reports involving 74 adverse events were received by the national PV unit - Increase in quality of ADR reports after refresher training | - Difficult to sustain ADR reporting and PV systems established - Underreporting of ADRs - Reporting affected by poor transport, access to health facilities, telecommunications and malaria transmission. | - PV focal point was key to the success of the implementation - Continuous training, feedback and supervisory visits essential to stimulate reporting - Means of transportation and communication is essential |
| Shrestha et al.,  Nepal  (37) | Educational | The main aim of the study was to assess the impact of an education intervention on the knowledge and attitude of HCP attached to the regional PV centre in an oncology based hospital of Nepal. | - 2h Training and interactive session on PV with distribution of posters and handouts | - Significant increase in mean knowledge score on PV and ADR as well as attitude towards PV. | None reported | - Collaboration of professionals and patients could drive and inform best practices in ADR reporting |
| SIAPS,  Swaziland  (38) | Passive and active safety surveillance systems | To introduce and implement the Sentinel Site-based active Surveillance System for antiretroviral and anti-TB (SSaSSa) treatment programs. | - Creation of the protocol and tools for the electronic SSASSA system - installed on 5 hospitals and one clinic - Development of a recruitment system at HIV and TB sites - Dissemination of PV data from the SSASSA system at both the national and regional levels." | - A total of 956 patients have been enrolled, and 58 adverse events have been recorded | - Data collection not optimal at all facilities | None reported |
| Siddiqui et al.,  India  (39) | Educational | To assess the knowledge, attitude, and practice of PV and ADR reporting of HCP before and after an educational intervention. | - Interactive educational session - Follow-up and reminders for one month | - Significant improvement in overall knowledge and attitude of HCP - ADRs increased from 0 to 10 during the 1 month follow-up period | - Underreporting of ADR due to a lack of knowledge and awareness and lack of time | None reported |
| Terblanche et al.,  South Africa  (40) | Mixed | To develop, implement and evaluate a structured pharmacist-driven PV system for in-patient ADR reporting. | - Retrospective review of ADR reports 18 months prior to the intervention - Needs analysis on structures, processes and outcomes of ADR reporting - ADR Training for HCP and support staff - Posters in wards and public areas to increase awareness - Increase access to ADR reporting forms and creating ADR reporting WhatsApp group - Supervisory visits by pharmacist - Distribution of SOPs on ADR reporting - Data analysis and feedback to HCP | - The number of HCP reporting any ADR significantly increased from 12.1% to 33.8% post the interventions. - Reasons for not reporting ADRs decreased significantly - HCP knowledge of the ADR reporting improved | - Unspecified operation challenges - The quality of the reports was not assessed | None reported |
| Tsafack et al.,  Cameroon  (41) | Mobile and electronic reporting | To assess the effect of telephone "beep" on community based reporting rates of AEFI during routine immunization sessions in a Cameroon Health District. | - Parents of vaccinated children were randomly assigned: i) to receive a phone contact from the investigation team and was advised to ''beep'' (short phone call not picked up) the investigators team in the case any medical incidence occurs within the 30 days following the immunization (intervention group) or; ii) to return to the health facility in case any medical incidence occurs within the same period (control group). | - The use of "beep" significantly increased ADR reporting rate compared to routine recommendations - 20 AEFI were reported within 30 days after vaccine administration; 19 of these in the intervention group and 1 AEFI in the control group. | - Limited number of participants - Selection bias - majority of participants resided in an urban zone and therefore more accustomed to the use of the telephone “beep” | - Mobile phone based tools like SMS have the capacity to complement existing passive reporting systems but the implication of a cost could be a barrier to reporting |
| UMC,  Mexico  (42) | Mobile and electronic reporting | To implement Vigiflow, the UMC-developed system for reporting ADRs in Mexico. | - The implementation process included workshops on using VigiFlow and VigiLyze for PV | - Adopting VigiFlow, enabled COFEPRIS to carry out its essential activities dynamically, with high-quality information. - Vigiflow and e-reporting have been implemented throughout the country, by all stakeholders | None reported | None reported |
| Varallo et al.,  Brazil  (43) | Educational | To assess the impact of a multifaceted educational intervention on the prevalence of ADR reporting. | - A lecture on PV concepts with a practical class on completing ADR reports - Distribution of educational materials | - 70-fold increase in the number of ADR reports during the 12 months post-intervention, though this decreased after the first 4 months - Improvement in form-completion skills PV knowledge | - PV training not included in new hospital risk management and patient safety policy | - The number of reports decreased after the first four months, suggesting the need for periodic retraining |
| Vo et al.,  Vietnam  (44) | Educational | The study aimed to describe ADR reports after an educational intervention for physicians and nurses by clinical pharmacists. | - Training of nurses and doctors on reporting of ADRs and diagnosis, treatment, and reporting of ADRs, respectively | - Significant increase in the quantity (12-fold compared to pre-intervention) and quality of ADRs reported | - Increase in reporting rate observed in the first 8 months of follow-up was not sustained | - More intensive and specific training and other measures are needed to improve under-reporting of ADRs |
| Vogler et al.,  Brazil  (45) | Mobile and electronic reporting | To describe steps taken by Anvisa to make Brazil compliant with international PV standards and to increase the number of ADRs collected. | - VigiFlow Setup - VigiFlow Operation - eReporting module, regional centre module and MAH module - Promotion and Training of HCP | - 62.6 % increase in ADRs received by Anvisa compared to the same period for the previous year | None reported | None reported |
| Zaveri et al.,  India  (46) | Educational | Impact of educational training on knowledge, attitude, and practice of pharmacovigilance in nursing staff of tertiary care hospital. | - One-hour interventional lecture on ADRs and PV in India with a practical session on how to complete an ADR reporting form | - Significant increase in knowledge and attitude towards PV however only 12% of participants reported an ADR | None reported | None reported |
| 2. Interventions aimed at strengthening various components of the national PV system | | | | | | |
| Adenuga et al.,  Namibia  (47) | Mixed | To explore the challenges associated with the effective integration of PV systems in public healthcare in a developing country such as Namibia. | - Systems-related ways of improving PV and ADR-reporting -make PV routine, implement electronic reporting - Promote advocacy and awareness of PV among HCP - Include PV into educational curriculum and continuous education for HCP - Incentivisation for PV activities including feedback to HCP and recognition of best performers - Inclusive reporting by providing user-friendly ways such as mobile phones - Stakeholder engagement | None reported | - Weak PV policies and structures - Limited capacity and support for implementation of PV activities - Negative attitude of healthcare workers towards PV | None reported |
| Akel et al.,  Lebanon  (48) | Mixed | Engage pharmacists in reporting the adverse drug reactions by creating an efficient tool for this purpose. | - Creating a Medication Safety Subcommittee - Designing reporting tools and the method of report analysis - Assessing field medication safety culture; training and continuing education on medication safety | - This was a pilot testing - the first 20 ADRs reported would enable Lebanon to become a member of WHO PIDM - Better knowledge and ADR awareness after training | None reported | - The study highlighted the need of educational programs to emphasize the role and responsibility of pharmacists in PV practices |
| Chakrabarty et al.,  India  (49) | Mixed | To inform, educate, and enlighten the readers about the constitution and dynamics of a PV centre. | - Setting up a PV centre with adequate logistic and infrastructure - Stakeholder communication and buy-in - Develop and disseminate ADR forms and PV tools - Ensure adequately qualified and trained staff - Acquiring a PV database - Foster a reporting culture - Establish procedures for PV tasks - Ensure stable funding | None reported | None reported | None reported |
| Diomandé et al.,  Benin, Burkina Faso, Cameroon, Chad, Ethiopia, Ghana, Niger, Nigeria, Senegal, Sudan, and The Gambia  (50) | Mixed | Establish vaccine PV systems in several African countries for post authorisation safety surveillance of for AEFI after PsA-TT mass immunization campaigns. | - Four-day training workshop - National safety Committees established and trained - National PV experts trained - Country and regional coordination teams designated and trained on their roles, and responsibilities - Active surveillance for specific AEFIs | - Implementation of national safety committees - Active surveillance of conditions of interest | - Resources allocated to AEFI monitoring decreased over time, negatively impacting data quality - Poor collaboration and coordination between NRA and EPI - Poor safety communication strategy | - Continuous global advocacy to mobilize more resources to build national vaccine PV systems - Coordination between the EPI and the NRA essential for planning and implementation of vaccine PV systems |
| Elshafie et al.,  LMIC  (51) | Mixed | To propose different strategies to encourage the introduction and sustain the advancement of robust pharmacovigilance systems in developing countries. | - Training HCP using different media and incorporation PV into educational institutions and curricula of HCP - Active public engagement in PV - Facilitate ADR reporting by simplifying the process and enabling mobile and electronic reporting - Adequate data management and communication of safety signals - Well-organized healthcare systems, with effective policies and strict drug regulations - Regulations and guidelines for industry - Curbing falsified drugs | None reported | None reported | None reported |
| Hartigan-go,  Philippines  (52) | Mixed | To strengthen the national PV system by reinforcing consumer reporting. | - Training courses on PV - Establish a national notification system - Advocacy and awareness initiatives - Introduction of a user-friendly report form - A simple causality assessment process was designed by an advisory committee - Feedback and appreciation letters and information about the reports submitted | - Over 1600 reports from various areas of the country received since - Health advisories and warnings to the health professionals and to the public - Medical device issues led to change in government procurement sources | - Underreporting of herbal medicines related ADRS | None reported |
| Hartigan-go,  Philippines  (53) | Mixed | To describe the establishment of the Philippine PV system and the lessons learnt. | - Education forum and training workshop for consumers - Simplified online (e-ADR) notification process - Distribution of advocacy materials such as posters and training materials | - No dramatic increase in consumer reporting observed - Quality and completeness of reports often insufficient for causality assessment - Regulatory actions such as advisories and product confiscation | None reported | - Consumers and patients are important allies in ensuring the safety and efficacy of medicines - Social media can be a useful tool to increase awareness and gather information about PV |
| Joshi et al.,  India  (54) | Mixed | To describe vaccine safety and surveillance for AEFI in India. | - Development of first operational guidelines in 2005 and revised in 2015 - Establishment of state and national AEFI committees - Appointment of four Zonal AEFI consultants - AEFI committees trained in investigation and causality assessment - Development of communication tools and guidelines - Implementation of a quality management system" | - Functional AEFI surveillance system | - The number of the reported serious AEFI are still far less than the expected numbers - Although the AEFI committees at the district and state levels have been established, a large proportion are far from functional | - AEFI surveillance program should be ready to assume greater responsibility comprehensively respond to the community concerns and sustain public confidence in vaccines |
| Jusot et al.,  Malawi  (55) | Mixed | To improve reporting of AEs by strengthening passive safety surveillance via PV training and mentoring of local PV stakeholders and HCP. | - Two-day training of EPI and NRA on PV - Abridged PV training for HCP - Regular PV mentoring on site or via phone calls - Logistic support to upgrade the national PV centre - computer system, PV database - Appointing a national PV coordinator, data manager and district PV focal points - Strengthening mechanisms for ADR reporting | - Regular PV training and mentoring of HCP were effective in enhancing passive safety surveillance in Malawi. - Significant increase of number of AEs reported | - Unavailability of a safety database during the implementation period - Transmission of reports to the national PV centre - No functional national safety committee during the pilot | - Continuous mentoring of HCP maintains motivation - Designating a national PV coordinator is important - Creation of partnerships and leveraging opportunities is essential for synergy |
| Kalaiselvan et al.,  India  (56) | Mixed | To describe measures taken by the National Coordination Center (NCC) for PV Programme of India (PvPI) to enhance patient safety including capacity building for monitoring, surveillance, collaboration with national health programs and other organizations to increase ADR reporting. | - Establishing a Culture of Adverse Drug Reaction Reporting - Integration of PV Programme of India and Public Health Programs (PHP) - Collaborations With Central Drugs Standard Control Organization - Education and Training on PV at Regional Training Centres - Safety communications via website, media, newsletter - Helpline Facility to Provide Assistance in Adverse Drug Reaction Reporting - Android Mobile Application for Adverse Drug Reaction Reporting - Utilization of Periodic Safety Update Reports Reporting - Availability of Medicine Side Effect Reporting Form for Consumers in Different vernacular languages | - The contribution of India to the WHO global Individual Case Safety Reports (ICSRs) database is 3% - ADR reporting through PvPI improved with the measures such as education, training, and provision of technical assistance | None reported | None reported |
| Meher,  India  (57) | Mixed | To describe vaccine safety and surveillance for AEFI in India | - Development of AEFI guidelines in 2005, revised in 2010 and 2015 - AEFI committees were set up in 2008 - National AEFI Secretariat and National AEFI Technical Collaborating Centre were established for technical oversight and support - Launch of the Pharmacovigilance Program of India (PvPI) in 2010 - Setting up ADR monitoring centres in different parts of India | None reported | None reported | None reported |
| Mehta et al.,  South Africa  (58) | Mixed | To outline findings and recommendations of a national pharmacovigilance workshop held in August 2012 in South Africa. | - Develop a national PV plan underpinned by five key principles: - Regulatory, programmatic and institutional PV incorporated into a cohesive national system - The national PV system should contribute to treatment policy decision-making and improved patient care - The national PV system should incorporate both passive and active surveillance methods and leverage successes - Investment in capacity building and training in PV and pharmacoepidemiology - Feedback and communication to stakeholders | None reported | None reported | None reported |
| NAFDAC,  Nigeria  (59) | Mixed | Presentation of the National Agency for Food and Drug Administration and Control (NAFDAC) Strategic Plan. | - Strategic focus 3: Safety and Quality of Regulated Products – to strengthen the Platform for Easy Reporting of Adverse Events and Timely Resolution | None reported | None reported | None reported |
| NDA,  Uganda (60) | Mixed | To provide a strategy to guide PV activities for the period 2018 to 2022. | - Enhance technical capacity at all levels involved in PV to improve safety data management and signal detection. - Strengthen existing reporting systems through implementation of smart safety surveillance initiatives. - Ensure that existing laws, regulations and guidelines comprehensively cover all areas of drug regulation including marketing authorisation holders. - Enhance information exchange between all PV stakeholders, both national and international, and facilitate collaboration through harmonisation of activities. | None reported | None reported | None reported |
| Nigeria MOH,  (61) | Mixed | To provide a framework and holistic approach for the implementation of the national PV policy in all tiers of the healthcare system. | - Setting up of PV structures - Establishing procedures to achieve defined outcomes and impacts by key PV partners - Define key performance indicators to measure performance and outputs | None reported | None reported | None reported |
| Nguyen et al.,  Vietnam  (62) | Mixed | To provide an overall picture of the Vietnamese PV system since the establishment of the National Drug Information and Adverse Drug Reaction Monitoring Centre (NDIADRMC) and describe lessons learnt in a resource-limited country when dealing with PV issues. | - NDIADRMC established in 2009 - Between 2010 and 2015, the Ministry of Health issued a number of legal documents related to PV - Linkages between PV activities and targeted PHPs have been established and improved over time - Education of HCP | - Functional safety surveillance system | - Variation in the quality and quantity of reports between regions, hospitals, and HCP - Limited Communication, Training, and Information Sharing - Divisions Between Activities Coordinating Public Health - Lack of Involvement from Private Drug Retailers, Customers, and Public Media | None reported |
| Nzolo et al.,  DRC  (63) | Mixed | To share the experience of the Democratic Republic of Congo (DRC) in developing its own PV System and to put its best practices and bottlenecks into a broader perspective. | - 5-day training of PV focal points, sensitization of HCP, establishment and training of NECs - Establishing reporting and feedback mechanism - Supervision visits - Enhanced spontaneous reporting at the community level - Active PV (cohort event monitoring) for specific drugs implemented | - Creation of national PV centre - Implementation of PV at Kinshasa university - Signal detection activities initiated in DRC - Collaboration established between PV stakeholders - VigiFlow™ installed and functional - PV included in curriculum of medicine, pharmacy, and dentistry schools | - Politics issues around location of national PV centre and conflict of interest between stakeholders - Lack of funding - Lack of comprehensive and validated guidelines for PV activities - Poor response from some stakeholders | - Improved stakeholder collaboration at all levels essential to incorporate PV into the national health system |
| PAVIA,  LMIC  (64) | Mixed | To provide the critical steps and actions that should be put in place in a proactive and well-coordinated manner in order to effectively implement a national pharmacovigilance policy. | - Stakeholders Engagement - Statutory Endorsement - Institutionalization of the Pharmacovigilance Policy - Addressing the Roles and Responsibilities of Stakeholders - Secure funding" | None reported | None reported | None reported |
| Sydykov et al.,  Republic of Kazakhstan  (65) | Mixed | To describe the key stages in the development of pharmacovigilance in the Republic of Kazakhstan. | - Creation of the national PV centre in 2002 - 2005 to 2008 - development of the system of ADR monitor - Establish regulatory framework for public PV system - 2009 improving and harmonizing the regulatory documents in drug safety control. | None reported | - Low activity of manufacturers on reporting of ADRs - Underreporting by HCP - Poor understanding and involvement of patients and consumers - Lack of an integrated information system for all participants in the monitoring of ADR | None reported |
| Tanani et al.,  Morocco  (66) | Mixed | To demonstrate the various steps of successful model of integrating PV in Moroccan Tuberculosis Control Program (PV-MTCP). | - Designation of national coordinator of PV - New technical committee on PV established - Collection of reports and feed back to reporters - Electronic recording of ADR reports - Data processing and signal management | - Significant increase in number of ADR reports (3.6 to 37.4 cases/month) after integration of the PV programme | - Patient files for those who have did not develop ADRs were unavailable to estimate the incidence of ADRs and risk factors | None reported |
| TDR,  Malawi  (67) | Mixed | To introduce innovative techniques to enhance reporting of adverse drug reactions and adverse events following immunization. | - A nationwide media campaign to raise awareness of the public and enhance reporting of suspected ADRs - Extensive cascade training targeting district-level PV focal points - Measure the impact of the training programme, via a Knowledge, Attitude and Practice survey - Developed a USSD (Unstructured Supplementary Service Data) platform for reporting ADRs and AEFI | - 2.8 fold increase in the rate of ADR detection and 1.8 fold increase in ADR reporting rate - Significant increase in ADR reports from 15 to 297 six months after the training | - Lack of reporting forms, delay in transferring reports and lack of feedback | None reported |
| TMDA, Tanzania (68) | Mixed | To develop a roadmap to outline activities to be undertaken to overcome gaps and challenges  identified during the baseline situational analysis. | - Improve the efficiency and functioning of regulatory and organizational structures of PV activities - Define and clarify the roles and responsibilities for all PV stakeholders - Increase the effectiveness of active (sentinel) surveillance of ADRs - Improve connectivity of databases and use of PV tools for event detection, reporting, analysis and dissemination - Increase resources to sufficiently exercise safety -monitoring - Improve PV-relevant skills and competencies at various levels - Improve monitoring and evaluation of the performance of the PV system - Enhance national, regional and international initiatives and   networking in relation to PV skills, knowledge and better resource mobilization and utilization | None reported | None reported | None reported |
| WHO  Malawi, Ghana,  Kenya  (69) | Mixed | To describe PV readiness indicators of vaccine PV in preparation for vaccine introduction. | - PV harmonization and capacity-building - Focused refresher training programmes and root cause analysis - Development of tools, guidelines, job aids - Establishment and training of national safety committees - Shared line-lists, printing and distribution of reporting tools and surveillance manual | - Vaccine safety surveillance systems strengthened - Increase reporting of AEFI | None reported | - The baseline surveillance systems established in these countries could be used for detecting AESI associated with other vaccine introductions, such as for COVID-19 |
| WHO  Global  (70) | Mixed | To propose a strategy to strengthen PV capacity in LMICs and, in the long-term, establish end-to end safety surveillance of products from their clinical development to the post market stages. | - Adopt a stepwise approach with an initial pilot for three new products (two medicines and one vaccine) - Leverage available resources from partners - Develop integrated plans that include key marketing authorization holders - Develop a holistic plan for PV - Collaboration with other ongoing initiatives - Build PV infrastructure progressively, moving from minimum to mid-range and advanced capacity." | - Significant increase in ADR reporting rates - Increased quality of reports - Increased capacity for signal detection and increased capacity to assess risk management | None reported | None reported |
| WHO  Global (71) | Mixed | The Global Vaccine Safety Blueprint (GVSB) provides strategies to establish systems that optimize the monitoring of vaccine safety profiles throughout their life-cycle; moving from a minimal and enhanced capacity concept to maturity levels | - To help LMICs to implement at least minimal capacity for vaccine safety activities - To enhance capacity for vaccine safety assessment in countries that introduce newly-developed vaccines - To establish a global vaccine safety support structure." | None reported | None reported | None reported |

### **Appendix VI: Articles excluded on full text**

| 1. **Duplicates not initially identified (n=2)** | |
| --- | --- |
| 1 | Bomene DN, Lula Y, Ntamabyaliro N, Engo A, Tona G. Development of Pharmacovigilance System in a Resource-Limited Country, the Experience of the Democratic Republic of Congo. Drug Safety. 2018;41(11):1120. |
| 2 | Sturkenboom M, Perez-Vilar S, Weibel D, Black S, Maure C, Castro JL, et al. Building Capacity for Active Surveillance of Vaccine Adverse Events in Low and Middle-Income Countries. Pharmacoepidemiology and Drug Safety. 2017;26:241-2. |
| 2. **Global recommendations (n=3)** | |
| 3 | Graham, J. E., Borda-Rodriguez, A., Huzair, F., & Zinck, E. (2012). Capacity for a global vaccine safety system: the perspective of national regulatory authorities. *Vaccine*, 30(33), 4953-4959. |
| 4 | Letourneau M, Wells G, Walop W, Duclos P. Improving global monitoring of vaccine safety: a survey of national centres participating in the WHO Programme for International Drug Monitoring. Drug Safety. 2008;31(5):389-98. |
| 5 | Pal, S. N., Olsson, S., & Brown, E. G. (2015). The monitoring medicines project: a multinational pharmacovigilance and public health project. *Drug Safety*, 38(4), 319-328. |
| 3. **Survey / systematic review (n=4)** | |
| 6 | Agoro OO, Kibira SW, Freeman JV, Fraser HSF. Barriers to the success of an electronic pharmacovigilance reporting system in Kenya: an evaluation three years post implementation. Journal of the American Medical Informatics Association. 2018;25(6):627-34. |
| 7 | Paudyal V, Al-Hamid A, Bowen M, Hadi MA, Hasan SS, Jalal Z, et al. Interventions to improve spontaneous adverse drug reaction reporting by healthcare professionals and patients: systematic review and meta-analysis. Expert Opinion on Drug Safety. 2020;19(9):1173-91 |
| 8 | Suku CK, Hill G, Sabblah G, Darko M, Muthuri G, Abwao E, et al. Experiences and Lessons From Implementing Cohort Event Monitoring Programmes for Antimalarials in Four African Countries: Results of a Questionnaire-Based Survey. Drug Safety. 2015;38(11):1115-26. |
| 9 | Zhao Y, Wang T, Li G, Sun S. Pharmacovigilance in China: development and challenges. International Journal of Clinical Pharmacy. 2018;40(4):823-31. |
| 4. **Developed country (n=5)** | |
| 10 | Baek HJ, Cho YS, Kim KS, Lee J, Kang HR, Suh DI. Multidisciplinary approach to improve spontaneous ADR reporting in the pediatric outpatient setting: a single-institute experience in Korea. Springerplus. 2016;5(1):1435. |
| 11 | Black S, Zuber PLF. Global trends and challenges in vaccine safety. Pediatric Health. 2009;3(4):329-35. |
| 12 | Cheema E, Almualem AA, Basudan AT, Salamatullah AAK, Radhwi SO, Alsehli AS. Assessing the impact of structured education on the knowledge of hospital pharmacists about adverse drug reactions and reporting methods in Saudi Arabia: an open-label randomised controlled trial. Drugs and Therapy Perspectives. 2019;35(6):296-300. |
| 13 | Kang DY, Ahn KM, Kang HR, Cho SH. Past, present, and future of pharmacovigilance in Korea. Asia Pacific Allergy. 2017;7(3):173-8. |
| 14 | Tabali M, Jeschke E, Bockelbrink A, Witt CM, Willich SN, Ostermann T, et al. Educational intervention to improve physician reporting of adverse drug reactions (ADRs) in a primary care setting in complementary and alternative medicine. Bmc Public Health. 2009;9:11. |
| 5. **Outcomes of interest not clearly defined (n=7)** | |
| 15 | AbuAlRub RF, Al‐Akour NA, Alatari NH. Perceptions of reporting practices and barriers to reporting incidents among registered nurses and physicians in accredited and nonaccredited Jordanian hospitals. Journal of Clinical Nursing (John Wiley & Sons, Inc). 2015;24(19-20):2973-82. |
| 16 | Ceballos, M., Salazar-Ospina, A., Sabater-Hernandez, D., & Amariles, P. (2020). Evaluation of the effects of a drug with fiscalized substance dispensation, health education, and pharmacovigilance continuing education program in Colombia drugstores and drugstores/pharmacies: study protocol of a multicenter, cluster-randomized controlled trial. *Trials [Electronic Resource]*, 21(1), 545. |
| 17 | Chahal, H. S., Kashfipour, F., Susko, M., Feachem, N. S., & Boyle, C. (2016). Establishing a regulatory value chain model: An innovative approach to strengthening medicines regulatory systems in resource-constrained settings. Pan American Journal of Public Health, 39(5), 299-305. |
| 18 | Lahariya C, Paruthi R, Bhattacharya M. How a New Health Intervention Affects the Health Systems? Learnings from Pentavalent Vaccine Introduction in India. Indian Journal of Pediatrics. 2016;83(4):294-9. |
| 19 | Patel, H., & Gurumurthy, P. (2019). Improving medication safety in oncology care: impact of clinical pharmacy interventions on optimizing patient safety. *International Journal of Clinical Pharmacy*, 41(4), 981-992. |
| 20 | Preston C, Valdez ML, Bond K. Strengthening Medical Product Regulation in Low- and Middle-Income Countries. PLoS Medicine. 2012;9(10). |
| 21 | Shankar PR, Humagain B, Piryani RM, Jha N, Osti B. Establishing and strengthening a medicine and therapeutics committee in a medical college in Nepal: initial experiences. Pharmacy World & Science. 2009;31(2):241-5. |
| 6. **PV system assessment (n=7)** | |
| 22 | Alshammari TM, Alenzi KA, Ata SI. National pharmacovigilance programs in Arab countries: A quantitative assessment study. Pharmacoepidemiology and Drug Safety. 2020;29(9):1001-1 |
| 23 | Alshammari TM, Mendi N, Alenzi KA, Alsowaida Y. Pharmacovigilance Systems in Arab Countries: Overview of 22 Arab Countries. Drug Safety. 2019;42(7):849-68. |
| 24 | Barry A, Olsson S, Minzi O, Bienvenu E, Makonnen E, Kamuhabwa A, et al. Comparative Assessment of the National Pharmacovigilance Systems in East Africa: Ethiopia, Kenya, Rwanda and Tanzania. Drug Safety. 2020:1-12. |
| 25 | Kabore L, Millet P, Fofana S, Berdai D, Adam C, Haramburu F. Pharmacovigilance systems in developing countries: an evaluative case study in Burkina Faso. Drug Safety. 2013;36(5):349-58. |
| 26 | Kaewpanukrungsi W, Anantachoti P. Performance assessment of the Thai National Center for Pharmacovigilance. International Journal of Risk & Safety in Medicine. 2015;27(4):225-37. |
| 27 | Olsson S, Pal SN, Stergachis A, Couper M. Pharmacovigilance activities in 55 low- and middle-income countries: a questionnaire-based analysis. Drug Safety. 2010;33(8):689-703. |
| 28 | Opadeyi, A. O., Fourrier-Reglat, A., & Isah, A. O. (2018). Assessment of the state of pharmacovigilance in the South-South zone of Nigeria using WHO pharmacovigilance indicators. *BMC Pharmacology & Toxicology*, 19(1), 27. |
| 7. **Articles in other languages (n=8)** | |
| 29 | Cai T, Zhan SY. Develop the active surveillance system for vaccine safety in China. Chung-Hua Yu Fang i Hsueh Tsa Chih [Chinese Journal of Preventive Medicine]. 2019;53(7):664-7. |
| 30 | Cao X, Tang L, Cai Z. Effects of clinical pharmacist intervention on the quality monitoring of adverse drug reactions. [Chinese]. Pharmaceutical Care and Research. 2015;15(3):221-3. |
| 31 | Koryanova KN, Matveev AV, Egorova EA, Bekirova EY. Features of International and Regional Pharmacovigilance Systems. Regionologiya-Regionology- Russ J Reg Stud. 2020;28(3):571-97. |
| 32 | Mira J.J. CM, Montserrat D., Rodriguez J., Santacruz J., . Key elements in implementing adverse event notification systems in Latin American hospitals. Rev Panam Salud Publica. 2013;33(1):7. |
| 33 | Pepe V. L. E. aNHMD. National Pharmacovigilance Systems in Brazil and Portugal: similarities, differences, and challenges. Cad Saúde Pública. 2020;36(7). |
| 34 | Sánchez I, Amador C, Plaza JC, Correa G, Amador R. Assessment of an active pharmacovigilance system carried out by a pharmacist. Revista medica de Chile. 2014;142(8):998-1005. |
| 35 | Soyalan M, Demirdamar R, Toklu HZ, Gumusel B. National pharmacovigilance system and current practice in the Turkish Republic of Northern Cyprus. Marmara Pharmaceutical Journal. 2012;16(3):159-63. |
| 36 | Trujillo JC, De Guzman MA. Pharmacovigilance: ''Vigilantia initiative''. Pharmaceuticals Policy and Law. 2016;18(1-4):157-62. |
| 8. **Full texts not found (n=19)** | |
| 37 | Amarnath S, Sharma A, Jaikumar S, Basalingappa S, Ramaswamy S, Thulasimani M. Impact of an educational intervention on the awareness of pharmacovigilance among pharmacy and nursing students in Puducherry. Research Journal of Pharmaceutical, Biological and Chemical Sciences. 2014;5(2):1130-6. |
| 38 | Bao J. Strengthening HIV Pharmacovigilance in China. Drug Safety. 2018;41(11):1115. |
| 39 | Caro-Rojas, A. (2018). How the Colombian Pharmacovigilance Association Join the Stakeholders in Pharmacovigilance, and Promote the Best Practices in Latam? (Meeting Abstract). Drug Safety, 41(11), 1114-1114. <Go to ISI>://WOS:000446470800031. |
| 40 | Copucho HC, Reis LV, Siquoira SM, Pereira LRL. Strategies Impact to Increase the Reporting of Pharmacovigilance in a Brazilian University Hospital. Drug Safety. 2008;31(10):953- |
| 41 | Dodoo ANO, Marfo FP. Effect of an educational intervention on improving drug safety awareness amongst sellers of non-prescription medicines in a developing country. Drug Safety. 2005;28(10):947- |
| 42 | Gilani Z, Koech DC, Mecca L, Khaemba C, Mandale M, Mwanje W, et al. Strengthening the vaccine safety system in Kenya: assessment of best practices for vaccine safety among healthcare workers in Kenya. American Journal of Tropical Medicine and Hygiene. 2019;101:428-. |
| 43 | Gossell-Williams M, Paul T. Introducing medical students to pharmacovigilance through a Basic Research Skills Special Study Module. International Journal of Risk and Safety in Medicine. 2020;31(2):81-7 |
| 44 | Gupta YK. Ensuring patient safety - launching the new pharmacovigilance programme of India. Pharma Times. 2010;42(8):21-6. |
| 45 | Jamekornkul C, Suwannakaesawong W, Uerchaikul C, Preechathaveekid S. Development of Pharmacovigilance Networking System in Thailand. Drug Safety. 2012;35(10):935. |
| 46 | Ndiaye Y. Community based pharmacovigilance, a way forward for strengthening the pharmacovigilance system in African underserved areas: experience in Saraya health district in rural Senegal. American Journal of Tropical Medicine and Hygiene. 2010;83(5):77-. |
| 47 | Nyakiba, J. O., McMillan, M., & Kenyatta, G. (2014). Reporting and documentation of adverse drug reactions by health care professionals at a Kenyan public hospital: A best practice implementation project. JBI Database of Systematic Reviews and Implementation Reports, 12(7), 521-533. doi:http://dx.doi.org/10.11124/jbisrir-2014-1362. |
| 48 | Okalebo FA, Oluka MN, Guantai AN. Learning by Doing: Implementation of Pharmacovigilance in the Clinical Setting in a National Referral Hospital in Kenya. Drug Safety. 2018;41(11):1109-10. |
| 49 | Osakwe AI, Elemuwa UG, Akinola BO, Suku CK. Strengthening Pharmacovigilance System Through Leveraging Resources from Public Health Programmes in Nigeria. Drug Safety. 2011;34(10):930-1. |
| 50 | Ramirez VH. Efforts Made by the University in Costa Rica to Promote Pharmacovigilance. Drug Safety. 2018;41(11):1114. |
| 51 | Sakin SA, Hossain AM, Mahmud SH, Ahmed SM. Effect of Educational Intervention on Perception of Adverse Drug Reaction Reporting Among Medical Practitioners. Mymensingh Medical Journal: MMJ. 2020;29(2):399-404 |
| 52 | Thurin N, Puello A, Haramburu F. Pharmacovigilance System Implementation in a Middle-Income Country: the Case of the Dominican Republic. Drug Safety. 2015;38(10):1032. |
| 53 | Wong A, Sandron CA. Increasing drug safety and ADR reporting in Latin America: The crucial role of poison information centres. Toxicology. 2001;164(1-3):8-9. |
| 54 | Yavo JC, Assi SB, Kamagate M, Amari AS, Kouame KE, Adou FM, et al. Implementation of a national pharmacovigilance system and drug risk management in a French-speaking African country, Cote d'Ivoire: review and Prospects. Fundamental & Clinical Pharmacology. 2018;32:62. |
| 55 | Yousef T, Abbas A, Ali T, Abdelghadir A. The Experience of using Khartoum Medicines Information Center (Khmic) as a Focal Point to Enhance Pharmacovigilance In Sudan. Drug Safety. 2018;41(11):1187-8. |
| 9. **No interventions/strategy described (n=40)** | |
| 56 | Abiri, O. T., & Johnson, W. C. (2019). Pharmacovigilance systems in resource-limited settings: an evaluative case study of Sierra Leone. Journal of pharmaceutical policy practice, 12(1), 13. |
| 57 | Adenuga BA, Kibuule D, Rennie TW. Optimizing spontaneous adverse drug reaction reporting in public healthcare setting in Namibia. Basic & Clinical Pharmacology & Toxicology. 2020;126(3):247-53. |
| 58 | Ahuja V, Sharma V. Training in Post-authorization Pharmacovigilance. Perspectives in Clinical Research. 2010;1(2):70-5. |
| 59 | Amarasinghe A, Black S, Bonhoeffer J, Carvalho SM, Dodoo A, Eskola J, et al. Effective vaccine safety systems in all countries: a challenge for more equitable access to immunization. Vaccine. 2013;31 Suppl 2:B108-14. |
| 60 | Andrade PHS, de Almeida ACB, Dos Santos AKS, Lobo IMF, da Silva FA, da Silva WB. Challenges to the consolidation of pharmacovigilance practices in Brazil: limitations of the hospital pharmacist. Therapeutic Advances in Drug Safety. 2020;11:2042098620933748. |
| 61 | Apte AA. Reporting of adverse events for marketed drugs: Need for strengthening safety database. Perspectives in Clinical Research. 2016;7(3):111-4. |
| 62 | Babigumira JB, Stergachis A, Choi HL, Dodoo A, Nwokike J, Garrison LP, Jr. A framework for assessing the economic value of pharmacovigilance in low- and middle-income countries. Drug Safety. 2014;37(3):127-34. |
| 63 | Black S, Zuber PLF. Global trends and challenges in vaccine safety. Pediatric Health. 2009;3(4):329-35. |
| 64 | Bonhoeffer J, Kohl K, Chen R, Duclos P, Heijbel H, Heininger U, et al. The Brighton Collaboration - Enhancing vaccine safety. Vaccine. 2004;22(15-16):2046. |
| 65 | Chen RT, Shimabukuro TT, Martin DB, Zuber PL, Weibel DM, Sturkenboom M. Enhancing Vaccine Safety Capacity Globally: A Lifecycle Perspective. American Journal of Preventive Medicine. 2015;49(6 Suppl 4):S364-76 |
| 66 | Dylan Fernandes S, Anoop NV, Castelino LJ, Narayana Charyulu R. A national approach to pharmacovigilance: The case of India as a growing hub of global clinical trials. Research In Social & Administrative Pharmacy. 2019;15(1):109-13 |
| 67 | Elshafie S, Zaghloul I, Roberti AM. Pharmacovigilance in developing countries (part I): importance and challenges. International Journal of Clinical Pharmacy. 2018;40(4):758-63. |
| 68 | Guillard-Maure, C., Elango, V., Black, S., Perez-Vilar, S., Castro, J. L., Bravo-Alcantara, P., et al. (2018). Operational lessons learned in conducting a multi-country collaboration for vaccine safety signal verification and hypothesis testing: The global vaccine safety multi country collaboration initiative. Vaccine, 36 (3), 355-362. |
| 69 | Hamid AA, Ibrahim MIM. A Systematic Scoping Review of the State of Pharmacovigilance and Governance in the MENA Region: Challenges and Opportunities. Pharmaceutical Medicine. 2017;31(6):437-54. |
| 70 | Hartigan-Go K, Roces MC, Habacon CA, Mansoor O, Shin S. Developing an immunization safety surveillance system in the Philippines. Bulletin of the World Health Organization. 2000;78(9):1166. |
| 71 | Helali, A. M., Iqbal, M. J., Islam, M. Z., & Haque, M. (2014). The evolving role of pharmacovigilance and drug safety: The way forward for Bangladesh (Review). International Journal of Pharmaceutical Research, 6(4), 31-37. |
| 72 | Hussain, R., & Hassali, M. A. (2019). Current status and future prospects of pharmacovigilance in Pakistan. Journal of Pharmaceutical Policy & Practice, 12, 14. doi:https://dx.doi.org/10.1186/s40545-019-0178-x. |
| 73 | Kabore L, Yameogo TM, Sombie I, Ouedraogo M, Fofana S, Berthe A, et al. Plaidoyer pour un renforcement du système de pharmacovigilance au Burkina Faso. Sante Publique (Vandoeuvre-Les-Nancey). 2017;29(6):921-5. |
| 74 | Khan Z, Karatas Y, Rahman H. Adverse drug reactions reporting in Turkey and barriers: an urgent need for pharmacovigilance education. Therapeutic Advances in Drug Safety. 2020;11:1-3 |
| 75 | Kochhar, S., Edwards, K. M., Ropero Alvarez, A. M., Moro, P. L., & Ortiz, J. R. (2019). Introduction of new vaccines for immunization in pregnancy - Programmatic, regulatory, safety and ethical considerations. Vaccine, 37(25), 3267-3277. |
| 76 | Kucuku M. Role of pharmacovigilance on vaccines control. Journal of Rural Medicine. 2012;7(1):42-5. |
| 77 | Lokesh RV, Adusumilli PK, Shashi B, Kalaiselvan V, Nath SG. Integration of pharmacy education and pharmacovigilance: scope and challenges in india. Indian Journal of Pharmaceutical Education and Research. 2019;53(2):202-7. |
| 78 | Mahajan, V., & Saini, S. S. (2014). Improving AEFI surveillance in India. Indian Pediatrics, 51(3), 233. |
| 79 | Mandal, S. C., & Mandal, M. (2017). Evolution of pharmacovigilance programme: Present status in India. *Pharma Times*, 49(5), 31-36. |
| 80 | Mehta, U., Durrheim, D., Mabuza, A., Blumberg, L., Allen, E., & Barnes, K. I. (2007). Malaria pharmacovigilance in Africa: lessons from a pilot project in Mpumalanga Province, South Africa. Drug Safety, 30(10), 899-910. |
| 81 | Meyer, J. C., Schellack, N., Stokes, J., Lancaster, R., Zeeman, H., Defty, D., et al. (2017). Ongoing Initiatives to Improve the Quality and Efficiency of Medicine Use within the Public Healthcare System in South Africa; A Preliminary Study. Frontiers in Pharmacology, 8, 751. |
| 82 | Moscou K, Kohler JC. Matching safety to access: global actors and pharmacogovernance in Kenya- a case study. Global Health. 2017;13(1):20. |
| 83 | Mulchandani R, Kakkar AK. Reporting of adverse drug reactions in India: A review of the current scenario, obstacles and possible solutions. International Journal of Risk & Safety in Medicine. 2019;30(1):33-44. |
| 84 | Oliveira PMN, Lignani LK, Conceicao DAD, Farias P, Takey PRG, Maia MLS, et al. Surveillance of adverse events following immunization in the late 2010s: an overview of the importance, tools, and challenges. Cadernos de Saude Publica. 2020;36Suppl 2(Suppl 2):e00182019. |
| 85 | Olowofela A, Fourrier-Reglat A, Isah AO. Pharmacovigilance in Nigeria: An Overview. Pharmaceutical Medicine. 2016;30(2):87-94. |
| 86 | Olsson S, Pal SN, Stergachis A, Couper M. Pharmacovigilance activities in 55 low- and middle-income countries: a questionnaire-based analysis. Drug Safety. 2010;33(8):689-703. |
| 87 | Palaian S, Mohamed Ibrahim MI, Mishra P. Development of pharmacovigilance training module for community pharmacists in Nepal: A focus group study. Archives of Pharmacy Practice. 2016;7(4):130-5. |
| 88 | Perez-Vilar S, Weibel D, Sturkenboom M, Black S, Maure C, Castro JL, et al. Enhancing global vaccine pharmacovigilance: Proof-of-concept study on aseptic meningitis and immune thrombocytopenic purpura following measles-mumps containing vaccination. Vaccine. 2018;36(3):347-54. |
| 89 | Prakongsai N, Pongchaidecha M. Using collaborative program between pharmacists and nurses to improve spontaneous adverse drug reaction reporting system in Thailand. Value in Health. 2010;13(7):A537-A |
| 90 | Rachlis B, Karwa R, Chema C, Pastakia S, Olsson S, Wools-Kaloustian K, et al. Targeted Spontaneous Reporting: Assessing Opportunities to Conduct Routine Pharmacovigilance for Antiretroviral Treatment on an International Scale. Drug Safety. 2016;39(10):959-76. |
| 91 | Shukla S, Gupta M, Pandit S, Thomson M, Shivhare A, Kalaiselvan V, et al. Implementation of adverse event reporting for medical devices, India. Bulletin of the World Health Organization. 2020;98(3):206-11. |
| 92 | Simha H, Marisarla S, Mujeebuddin CS. Pharmacovigilance system in India. International Journal of Pharmaceutical Sciences Review and Research. 2020;63(1):73-80. |
| 93 | Stefanov O, Sharayeva M, Jajtchenja V. Development of pharmacovigilance system in Ukraine: first results. Pharmacoepidemiology & Drug Safety. 2004;13(3):197-9. |
| 94 | Varallo FR, Forgerini M, Herdeiro MT, de Carvalho Mastroianni P. Harmonization of Pharmacovigilance Regulation in Brazil: Opportunities to Improve Risk Communication. Clinical Therapeutics. 2019;41(3):598-603. |
| 95 | Zhou Y, Miller V, Hogan M, Callahan L. An overview of adverse drug reaction monitoring in China. International Journal of Pharmaceutical Medicine. 2006;20(2):79-85. |

### **Appendix VII: References**

21.	Kadima NJ, Nyiranteziryayo R, Umumararungu T, Adedeji AA. Use of mobile phones for patient self-reporting adverse drug reactions: A pilot study at a tertiary hospital in Rwanda. Health and Technology. 2020.

29.	Nkonde K, J.C. M, M.D. M, R.S. S. Implementation of interventions to strengthen the pharmacovigilance surveillance system at Dr George Mukhari Academic Hospital, Gauteng Province (Doctoral dissertation, Sefako Makgatho Health Sciences University) 2017.

38.	MSH. Strengthening Pharmacovigilance Systems in Swaziland to Improve Patient Safety and Treatment Outcomes. The Systems for Improved Access to Pharmaceuticals and Services (SIAPS) program; 2015.

60.	NDA. Monitoring safety of medicines in Uganda: strategies for better pharmacovigilance systems in settings with limited resources 2020.

61.	Health Mo. Nigerian national pharmacovigilance policy and implementation framework. 2020.

62.	Nguyen KD, Nguyen PT, Nguyen HA, Roussin A, Montastruc JL, Bagheri H, et al. Overview of Pharmacovigilance System in Vietnam: Lessons Learned in a Resource-Restricted Country. Drug Safety. 2018;41(2):151-9.

63.	Nzolo D, Kuemmerle A, Lula Y, Ntamabyaliro N, Engo A, Mvete B, et al. Development of a pharmacovigilance system in a resource-limited country: the experience of the Democratic Republic of Congo. Therapeutic Advances in Drug Safety. 2019;10:2042098619864853.

64.	PAVIA. PAVIA Guide for Effective Implementation of the Pharmacovigilance Policy in Resource Limited Settings (2021) PAVIA-EDCTP sponsored Project. . 2021.

### **Appendix VII: Abbreviations**

| **ADR** |  | Adverse drug reaction |
| --- | --- | --- |
| **AE** |  | Adverse events |
| **AEFI** |  | Adverse events following immunization |
| **AESI** |  | Adverse events of special interest |
| **ARV** |  | Antiretroviral |
| **BMGF** |  | Bill and Melinda Gates Foundation |
| **CADTH** |  | Canadian Agency for Drugs and Technologies in Health |
| **CINAHL** |  | Cumulative Index to Nursing and Allied Health Literature |
| **CHW** |  | Community health workers |
| **EPI** |  | Expanded Program on Immunization |
| **DRC** |  | Democratic Republic of the Congo |
| **GVSB** |  | Global Vaccine Safety Blueprint |
| **HCP** |  | Healthcare professionals |
| **JBI** |  | Joanna Briggs Institute |
| **JBI SUMARI** |  | JBI System for the Unified Management, Assessment and Review of Information |
| **LMIC** |  | Low-and-middle-income countries |
| **MOH** |  | Ministry of Health |
| **MSH** |  | Management Sciences for Health |
| **NAFDAC** |  | The National Agency for Food and Drug Administration and Control |
| **NDA** |  | National Drug Authority |
| **NDIADRMC** |  | National Drug Information and Adverse Drug Reaction Monitoring Centre |
| **NRA** |  | National regulatory authorities |
| **PAVIA** |  | PhArmacoVigilance Africa |
| **PHP** |  | Public health programs |
| **PIDM** |  | Program for International Drug Monitoring |
| **PRISMA_ScR** |  | Preferred Reporting Items for Systematic Reviews and Meta-Analyses Extension for Scoping Reviews |
| **PV** |  | Pharmacovigilance |
| **SIAPS** |  | Systems for Improved Access to Pharmaceuticals and Services Program |
| **SMS** |  | Short Message Service |
| **SSA** |  | sub-Saharan Africa |
| **SPS** |  | Strengthening Pharmaceutical Systems Program |
| **SwissTPH** |  | Swiss Tropical and Public Health Institute |
| **TMDA** |  | Tanzania Medicines and Medical Devices Authority |
| **TDR** |  | Special Programme for Research and Training in Tropical Diseases |
| **UMC** |  | Uppsala Monitoring Centre |
| **WHO** |  | World Health Organisation |
| **WHO GBT** |  | World Health Organisation Global Benchmarking Tool |
